# Geographically weighted Machine Learning for Spatial Prediction of Cancer Prevalence in the United States: A Mixed Method Approach

**DOI:** 10.64898/2026.08.18.26360598

**Authors:** Fereshteh Sadeghi Naieni Fard, Joseph R. Oppong, Chetan Tiwari, Kwadwo Boakye, Fariba Fard

## Abstract

Cancer prevalence is distributed unevenly across regions and caused by the interaction of multiple risk factors. Previous studies focused on the use of global modeling techniques to predict cancer at the county level that overlooks important spatial differences. This study aims to develop geographically weighted machine learning models to predict cancer prevalence at the census tract level in the United States and identify local determinants of cancer burden.

First, a scoping review was conducted to find a list of measurable drivers of cancer in the United States. Using this list, the data of these variables for 84415 census tracts were obtained from the Center for Disease Control and Prevention PLACES dataset and other publicly accessible resources. Then, several predictive models, including Ordinary Least Squares (OLS) and Geographically Weighted Regression (GWR), as well as Random Forest, XGBoost, and Deep Neural Network and their geographically weighted counterparts, were developed and compared using the Coefficient of Determination, Root Mean Square Error, and Absolute Error.

Results presented that geographically weighted models outperformed other methods, and geographically weighted XGBoost achieved the strongest and most consistent overall performance with pseudo-R2 ranging between 0.89 and 0.98. Feature importance analysis of this model illustrated that most important cancer drivers changed location by location. Aged people, racial composition, preventative behaviors, and metabolic conditions such as diabetes, hypertension, and high cholesterol were determined as influential predictors, although their relative importance varied across regions. These findings revealed the value of localized models at a small geographic scale to identify regional cancer risk patterns and help the allocation of proper resources to hotspot areas.

## Introduction

Cancer is a major cause of morbidity and mortality in the United States with 141.5 deaths per 100,000 population in 2023 (Center for Disease Control and Prevention [CDC], 2025). The number of cancer cases is expected to increase by 50 percent by 2030 and the national costs projected to reach 208.9$ billion (Islami et al., 2022; Ma et al., 2022; Gallicchio et al., 2022). Cancer is a multifactorial disease resulting from the interaction of multiple risk factors, and its prevalence has meaningful differences across regions (Obeng-Gyasi et al., 2021; Ma et al., 2022; Sobti et al., 2024; Islam et al., 2025).

Geographic location plays key role in forming environmental, social, and structural conditions. These geographic differences are caused by cultural norms, socioeconomic factors, and local policies as well as the inherent environmental features. These factors can also contribute to lifestyle changes and health behaviors (Buck, 2016; Sobti et al., 2024; Anderson et al., 2023). As a result, certain populations are more likely to experience poverty, and racism, and have limited access to healthcare services, and proper educational or food resources. They also have more exposure to environmental changes (Islami et al., 2022; Fletcher et al., 2020) which increase the risk of cancer development and prevent early diagnosis or timely treatment (Tesfaw & Muluneh, 2020; Fletcher et al., 2020; Salmeron et al., 2021; Islam et al., 2025).

In this context, accurate geographic prediction of cancer cases is a paramount step in preparing public health systems by reporting where cancer occurs and why. Using analysis at smaller geographic scales help to discover patterns ignored at larger levels (Tailor et al., 2019; Lynch et al al., 2020; Tabassum et al., 2025). These predictions can signal the real needs of vulnerable populations for resources and interventions before the conditions get worse (Rahib et al., 2021; Bray & Møller, 2006). The role of these predictions becomes more evident under crisis situations such as COVID-19 pandemic with dual pressure on cancer patients and healthcare systems (Robilotti et al., 2020).

Most previous studies analyzing geographic differences in specific types of cancer incidence, mortality, or screening across the United States used data at the level of county. For example, Ma et al. (2022) reported the substantial disparities of gastrointestinal cancer at the county level based on the modifiable and non-modifiable factors. With focus on environmental factors, Lee et al. (2024) presented the regional differences of lung cancer incidences at this level caused by environmental exposures. In another study Ahmed et al. (2021) predicted the mortality of lung and bronchus cancer at the county level using different predictors among which poverty and elevation were the most effective. Similarly, Palliyaguru et al. (2024) used different models to forecast the main risk factors of colorectal cancer and reported that low physical activity and obesity were significantly associated with this type of cancer in rural and urban counties. Luberice et al. (2025) also used the county level data of pancreatic cancer and illustrated how social determinants could shape the unequal distribution of the incidence rates of this disease. By including all types of cancer in the analysis, Ray & Ghosh. (2025) found that county level socioeconomic factors are the major causes of the differences in cancer mortality.

As such, most of these studies focused on only some particular types of cancer at the county level. Even studies conducted by Niu et al. (2022) and Bhattacharya et al. (2024) that introduced social determinants of cancer mortality at the census tract level, could find the determinants in the whole U.S without predicting hot spot areas or showing how the impact of these risk factors is changed location by location.

Also, previous studies used different methods in predicting cancer cases. Some of them applied linear regression methods such as the work done by Ma et al. (2022), and some used more advanced machine learning models including Random Forests, Gradient Boosting, and Bayesian Additive Regression Trees (Ahmed et al., 2021; Lee et al., 2024; Niu et al., 2022; Li et al., 2022; Dong et al., 2022) to model the nonlinear interactions between predictors. With more focus on finding the local and spatial differences, several studies have introduced geographically weighted models (Anderson et al., 2023; Gu et al., 2023; Shaik, 2024).

Despite these methodological advances, most of these studies used county level data for prediction of cancer cases. Moreover, geographically weighted machine learning models presented higher capabilities in identifying spatial differences and can significantly increase the accuracy of prediction which were not widely used in cancer research studies. Additionally, modeling of cancer cases needs to integrate and evaluate the interactions of all the risk factors which were ignored in previous studies. Addressing these gaps is very important to develop an accurate model that can be generalized and used in other geographic regions.

Accordingly, the present study is guided by several research questions. First, which factors are associated with spatial differences in cancer prevalence at the census tract level? Second, do geographically weighted machine learning models improve the predictive performance in comparison to global learners? Third, among the different models considered, which predicts cancer prevalence most accurately? Finally, to what extent does neighborhood-level modeling improve the identification of high-risk communities compared with county-level analysis?

To address these questions, this study first provides a comprehensive list of risk factors that can be measured at the census tract level. Using this list and after collection and integration of all variables, various geographically weighted machine learning models were developed, and their performance were compared with global learners. Then, according to the most accurate model, the contributing risk factors were determined at the census tract level. Results of this study increase the resiliency of the public health systems by providing appropriate methodology to predict cancer cases at small geographic levels and identify major risk factors contributing to increased cancer risk at this level.

## Methodology

The research methodology in this study contains four sequential phases. First, a collection of risk factors was created by conducting a scoping review. Using the results of the scoping review, relevant data was collected, preprocessed and integrated into a unique dataset at the census tract level. Then, geospatial adaptive models besides their global peers were developed using the curated dataset. Finally, the performance of those models was compared by applying different metrics to determine the best approach. Figure 1 displays the summary of research design for this study.

**Figure 1.**
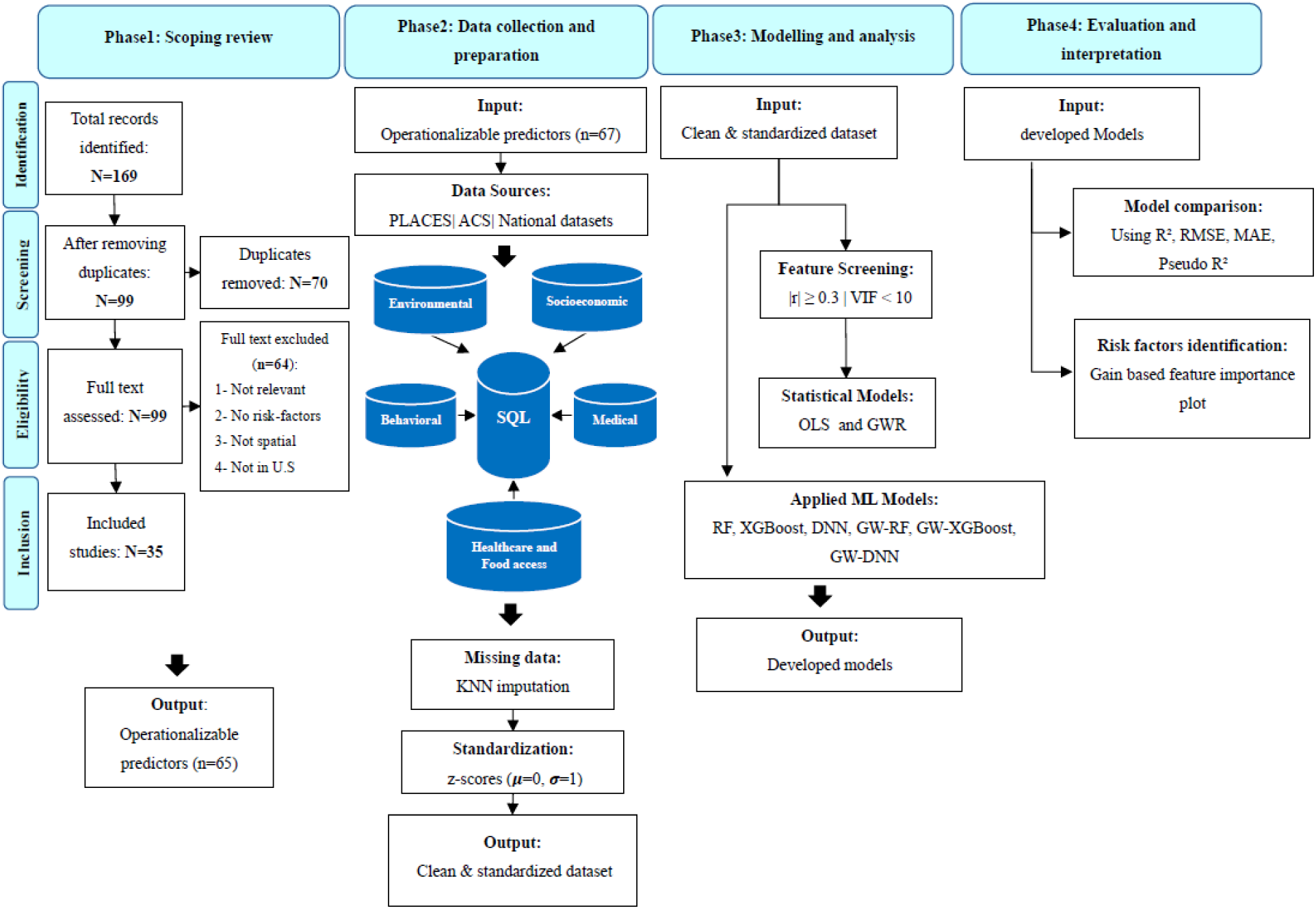
Summary of Research Design.

### Identifying Relevant Risk Factors: A Scoping Review

We examined available geospatial studies to understand which risk factors can affect cancer cases in the United States. This search was conducted using PubMed, Science direct, Scopus, and Google scholar. To include the most recent works, we focused on peer-reviewed studies published between 2016 and 2025, and we explored a broad range of search terms using combination of keywords such as “cancer”, “risk factor”, “determinant“, ”incidence“, ”prevalence“, ”mortality“, ”machine learning“, ”predictive modeling“, and ”analysis”. Specific keywords related to geography such as “county”, “state”, “zip codes”, and “census tract” were also incorporated into the search query. Only studies performed in the United States were included to comply with U. S. policies and data availability. The eligibility process began with screening of titles and abstracts. Studies that met the initial criteria then underwent full text review to find measurable variables associated with cancer risk.

### Data Collection and Preprocessing

Data corresponds to cancer prevalence rates in 2023, and several behavioral and clinical factors were obtained from the Center for Disease and Control (CDC) PLACES website. Data of other socioeconomic, environmental, healthcare and food access variables studied by 35 peer-reviewed papers were collected from publicly accessible U.S. sources, including the U.S. Census Bureau, Environmental Protection Agency (EPA), Agency for Toxic Substances and Disease Registry (ATSDR), National Aeronautics and Space Administration (NASA), National Centers for Environmental Information (NCEI), U.S. Geological Survey (USGS), U.S. Department of Agriculture (USDA), Food and Drug Administration (FDA), and the Inter-university Consortium for Political and Social Research (ICPSR).

Data of Artificial Light at Night (ALAN) were also extracted from satellite nighttime light image obtained from the Visible Infrared Imaging Radiometer Suite (VIIRS) in Google Earth Engine (GEE). We used a bilinear interpolation method (Sotomayor et al., 2023) to extract raster values at centroid of each census tract. Then, the level of exposure was calculated by measuring the average of the four closest pixels.

For some clinical factors including HIV and Hepatitis, prevalence rates were available at the county level. Following Hallisey et al. (2017), we assumed that within each county their rates are equally distributed, and their corresponding rate for each census tract were calculated using their population density. After this collection process, the final dataset contained 65 independent variables. All predictors were joined and integrated into a unique dataset using Federal Information Processing Standard (FIPS). See Appendix A that provides a detailed summary of the data sources, predictor variables, and their corresponding years of collection.

All states of the U.S has been included in the final dataset except five territories: American Samoa, Guam, the Northern Mariana Islands, Puerto Rico, and the U.S. Virgin Islands. Among these, 798 tracts had zero population. Hence, the values for other population-based predictors such as demographic variables and medical factors at these tracts were set to zero. Then, other missing values were imputed by using a k-nearest neighbors (KNN) method.

### Model Development

In this study two groups of models were developed and evaluated. The first group Consisted of Geographically weighted machine learning models including Random Forest, XGBoost and Deep Neural Network, using two approaches to design them. Then, the performance and generalizability of these models were compared with the second group, global models.

#### Geographically weighted ensemble models

To build Geographically Weighted Random Forest (GW-RF) and Geographically Weighted XGBoost (GW-XGBoost), first the United States is divided into 100 distinct spatial clusters using census tract centroids that each cluster contains a reasonable number of observations for developing local machine learning models. Then, 80 percent of all tracts were assigned to the training set and the remaining to the test set. Subsequently, local Random Forest (RF) and XGBoost models were developed in those clusters. Training samples in each cluster were selected due to their geographic proximity using a Ball Tree with haversine distance within the optimal bandwidth (Dolatshah et al., 2015). If the number of selected samples were insufficient, the k-nearest neighbors’ method was used to increase number of samples to reach optimum sample size obtained by evaluating Moran’s I spatial autocorrelation coefficient.

#### Geographically Weighted Deep Neural Network

This study followed the model proposed by Sun et al. (2021), in which the spatial distribution of downed dead wood volume in forest ecosystems in China was modeled. For each census tract i, the target variable *y_i_* presents crude cancer prevalence and is modeled as a function of a vector of standardized predictors 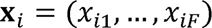 and geographic coordinates 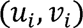 The intercept term is also incorporated, and an augmented feature vector was defined as 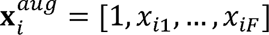, and the constant value is estimated as a baseline for the cancer prevalence which is independent of predictors and can be changed geographically.

This model has two subnetworks performed in parallel. The first network, Alpha branch, learns the measure of connection between predictors and cancer prevalence at each census tract. Alpha Branch generates coefficients specific to each geographic location using local predictor values and it can capture the nonlinear relationships that spatially changed between the cancer prevalence and risk factors. Alpha branch consists of multilayer perceptron with two hidden layers that have 64 neurons each. It uses rectified linear unit (ReLU) activation function with 0.4 dropout rate to avoid overfitting. the Alpha branch, denoted by *g_α_*(·), takes the input features and produces a set of local regression coefficients for each sample, 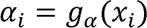

The second subnetwork which performs in parallel is named Spatial Weight Neural Network (SWNN) produces spatial weights of geographic locations. This subnetwork contains two hidden layers with 128 neurons each. It applied ReLU activation functions followed by a drop out layer with rate 0.4. The output layer uses SoftPlus activation function to generate nonnegative geographic weights. The SWNN, denoted as *g_w_*(·), takes the geographic coordinates, and produces spatial weights, . Equation (1) shows how the prediction is made by adding the input features, each multiplied by a geographic coefficient and a spatial weight where j shows the index of augmented predictors.

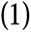

The model is trained by using the MAE loss function, and Adaptive Moment Estimation (Adam) optimizer at a learning rate of . It is trained for up to 100 epochs and with a batch size of 64. Figure 2 presents the architecture of the Geographically Weighted Deep Neural Network (GW-DNN). denotes for actual crude cancer prevalence and denote the predicted value for census tract .

**Figure 2.**
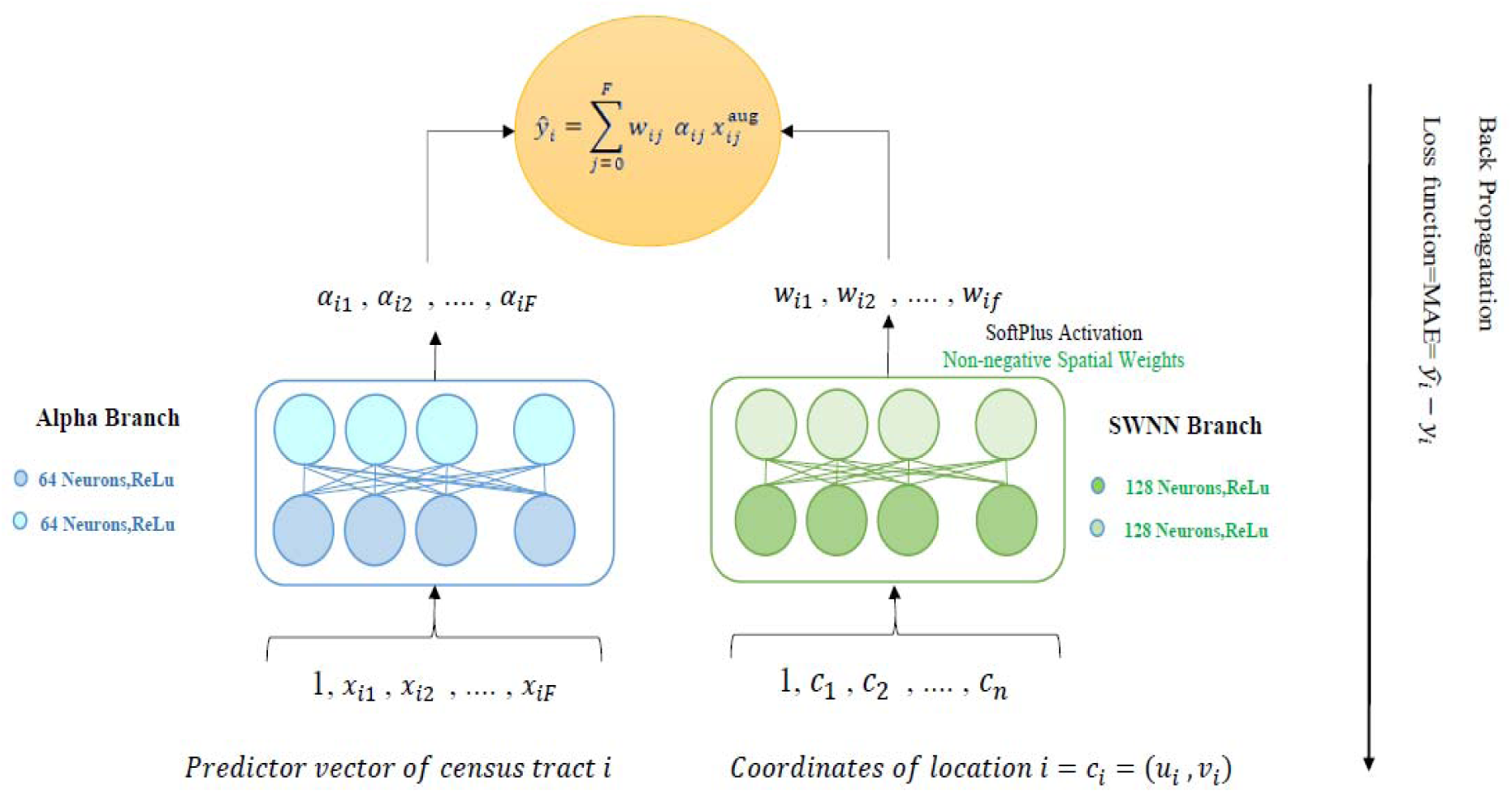
Geographically Weighted Deep Neural Network Model Architecture

### Feature Assessment and Parameters of the Model

This study used multiple steps for parameters selection. First, K-nearest neighbors (KNN) method is used to impute missing values. The optimal value of K was defined by using Moran’s I spatial autocorrelation. Although K=2 led to highest spatial autocorrelation with value 0.786, it includes only a few neighboring observations at the census tract level. Therefore, K=20 was selected because it provided a reasonable number of neighbors and still has strong spatial autocorrelation (Moran’s I > 0.675).

One of the critical parameters that needed to be defined in this study was the optimal bandwidth for both model development and selecting training and testing sets, and it was determined using cross validation on a stratified subsample of census tracts (n = 12,456; 40 tracts per state). The candidate range for bandwidth selection was between 500 to 1000 based on the study conducted by Ruckthongsook et al. (2018), which provides enough geographic details with reliable results. The final optimal bandwidth determined to be 500 according to the minimum corrected Akaike Information Criterion (AICc) value.

In addition to ensemble machine learning and DNN models, Ordinary Least Squares (OLS) and Geographically Weighted Regression (GWR) models were also developed in this study. These approaches are sensitive to multiclonality among variables and making feature assessment essential before model development. Hence, pairwise correlations were examined among all candidate predictors to evaluate potential multicollinearity. Figure 3 indicates the correlation heatmap of the predictors and the presence of correlations among several variables.

**Figure 3.**
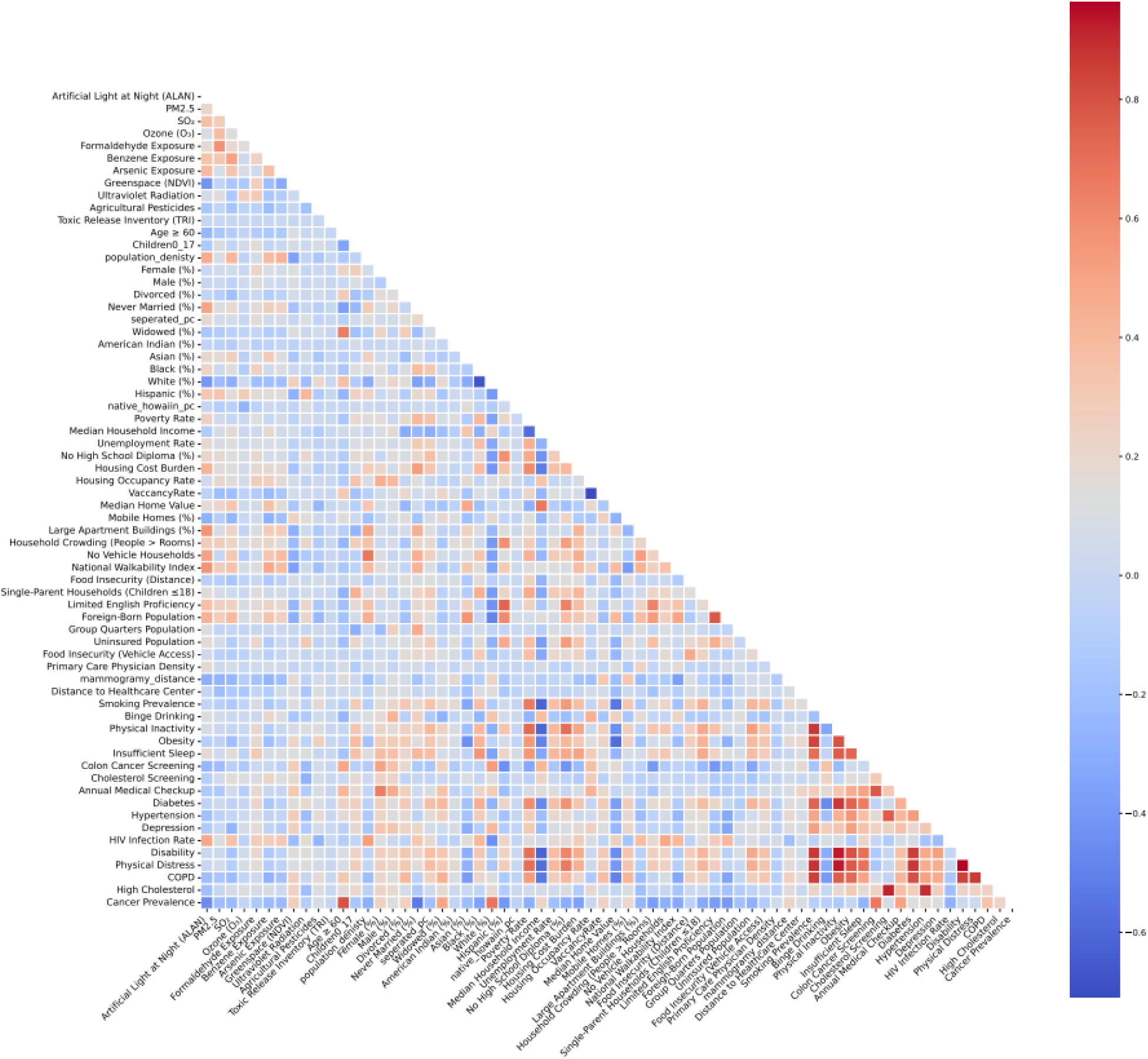
Correlation Heatmap of the Studied Variables

So, using two step variable selection procedure, the predictors with absolute Pearson correlation coefficient were kept for further analysis. Then, multicollinearity among these variables was evaluated using the variance inflation factor (VIF). Based on the literature, variables with VIF values below 10 were considered acceptable (O’brien, 2007; Vatcheva et al., 2016). Table 1 presented the final set of predictors.

**Table 1.** Candidate Feature Coefficients and their VIF Values.

| **Feature** | **VIF** | **Absolute Pearson's $r$** |
| --- | --- | --- |
| Colon cancer screening (%) | 7.52 | 0.672 |
| White (%) | 7.31 | 0.684 |
| Annual Medical Checkup (%) | 6.45 | 0.51 |
| Black (%) | 5.64 | 0.358 |
| Foreign-Born (%) | 4.43 | 0.391 |
| Limited English (%) | 4.12 | 0.384 |
| COPD Prevalence (%) | 3.01 | 0.348 |
| Hispanic (%) | 2.72 | 0.410 |
| Age $\geq 60$ (%) | 2.62 | 0.798 |
| Housing Cost Burden | 2.15 | 0.360 |
| Widowed (%) | 1.98 | 0.497 |
| Never Married (%) | 1.97 | 0.529 |
| Crowding | 1.91 | 0.378 |
| ALAN | 1.88 | 0.454 |
| Single-Parent Households<br>(Children $\leq 18$) (%) | 1.64 | 0.323 |
| National Walkability Index | 1.953 | 0.320 |

### Model Evaluation

For all ensemble and DNN models, the dataset was divided into training and testing subsets with 80 percent of the observations allocated to the training set and 20 percent to the test set. Then, the model performance was evaluated on both datasets to help compare the behavior of each model. Several evaluation metrics were used including the Coefficient of Determination (R^2^) measures the amount of the variance in target variable explained by the model, Root Mean Square Error (RMSE) evaluates the average magnitude of prediction errors and Mean Absolute Error measures the average absolute difference between predicted and observed values.

### Feature Importance Analysis

This study aimed to find the underlying factors associated with increased prevalence of cancer cases and to interpret the ability of the best-performing model to identify these factors. For the ensemble models, the feature importance method was employed (Alsahaf et al., 2024) which measures the contribution of each predictor to the performance of the model and how much it reduces the loss function in all decision trees.

To analyze the role of geographic locations in feature importance, the whole United States was divided into eight major regions, and separate feature importance analysis were conducted for each region to diagnose the most influential drivers. Besides that, three representative census tracts were selected to show more detailed analysis on how importance of the features changed across different geographic locations. See Appendix B for the table of eight major divisions of the United States.

## Results

According to the data preprocessing, the final dataset contains 71,255 census tracts with nonzero population. The mean of adult cancer prevalence rate is 7.91 percent with 2.82 standard deviation and Interquartile Range (IQR) of 8. Cancer prevalence rate ranged from minimum of 0.5 percent in census tract 503 located in Lubbock County, Texas to a maximum of 27.5 percent in census tract 31601 in Fairfax County, Virginia. About 95 percent of the census tracts presented prevalence values between 2.4 percent and 13.6 percent that indicated significant spatial differences across U.S. census tracts.

### Spatial distribution of cancer prevalence

The spatial prevalence of cancer cases in all census tracts showed meaningful differences in various regions (Figure 4). Several regions including parts of Appalachia, the Midwest, the Gulf Coast, and portions of the Northeast presented higher prevalence rate. Some selected areas in the Southwest and Pacific Northeast also display increased cancer prevalence. In contrast, lower prevalence rates are more common in some parts of Great Plains, Mountain West, and coastal regions. These patterns revealed that distribution of cancer cases are influenced by regional factors.

**Figure 4.**
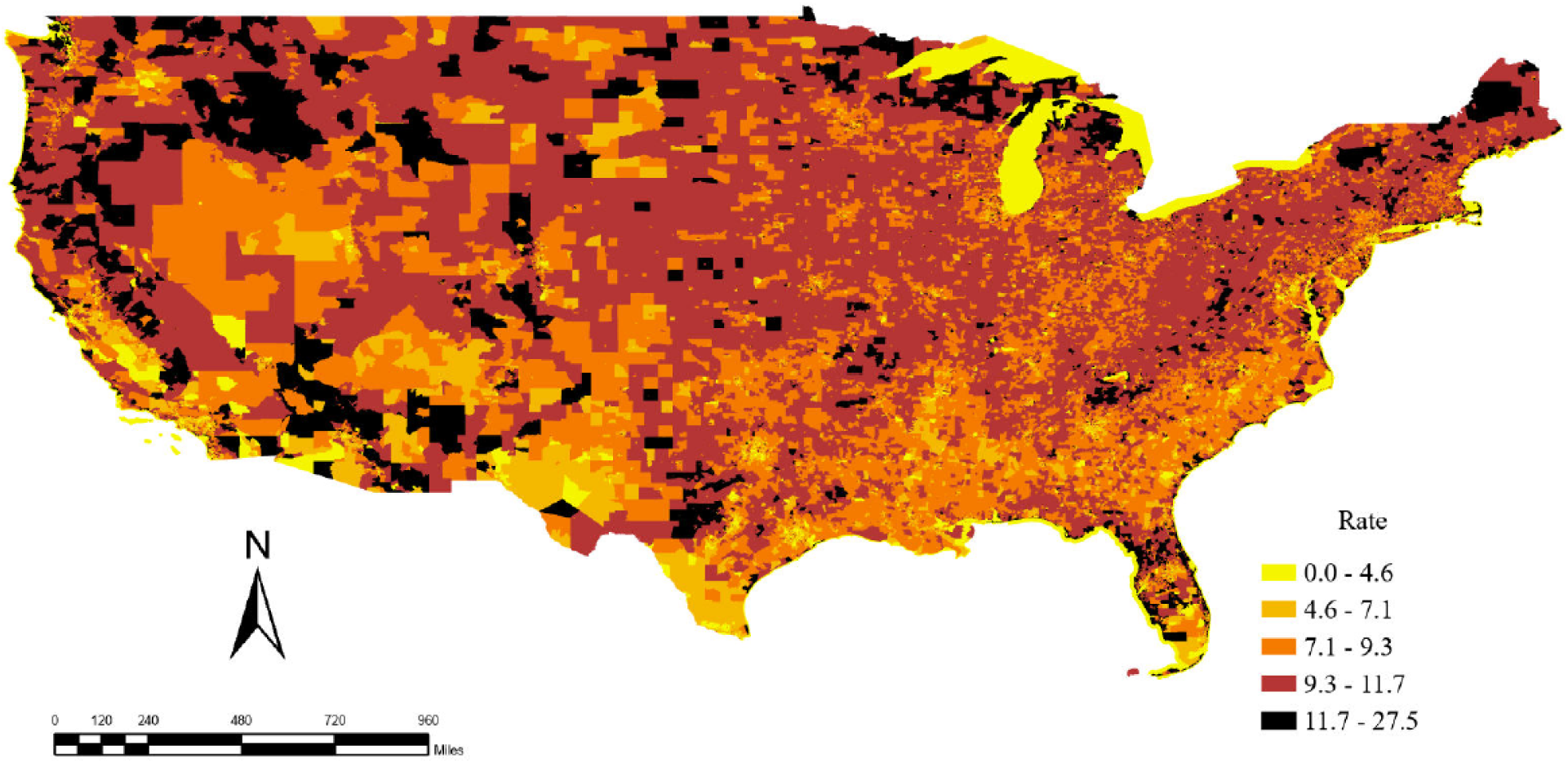
Census tract–level cancer prevalence rates in the United States (2023)

### Model Performance

Comparing the model performance of different predictive models presents the advantages of geographically weighted models. Among the global learners, XGBoost demonstrated the strongest predictive performance with a R² of 0.9840 on the training set and 0.9734 on the test set besides lowest MAE and RMSE. In addition, all geographically weighted models outperform their global counterparts. Among these, GW-XGBoost and GW-DNN model achieved the best overall performance. These performance comparisons illustrate that spatial non-stationary plays an important role in explaining the disparities in prevalence of cancer at the census tract level. Table 2 and Table 3 summarize the performance metrics for all models.

**Table 2.** Performance of Statistical Spatial Models.

| **Model** | **R<sup>2</sup>** | **RMSE** | **MAE** |
| --- | --- | --- | --- |
| **OLS** | 0.8985 | 0.8919 | 0.6687 |
| **GWR** | 0.9527 | 0.6085 | 0.4583 |

Note. This table presents the performance of metrics for statistical models including OLS and GWR

**Table 3.** Predictive Performance of Global Learners and Geographically Weighted Models.

| Model | Train $R^2$ | Train RMSE | Train MAE | Test $R^2$ | Test RMSE | Test MAE |
| --- | --- | --- | --- | --- | --- | --- |
| RF | 0.9858 | 0.3333 | 0.2325 | 0.9543 | 0.5935 | 0.4372 |
| GW-RF | 0.9958 | 0.1819 | 0.1343 | 0.9661 | 0.5108 | 0.3777 |
| XGBoost | 0.9814 | 0.3821 | 0.2953 | 0.9734 | 0.4525 | 0.3416 |
| GW-XGBoost | 0.9970 | 0.1528 | 0.1117 | 0.9802 | 0.3902 | 0.2887 |
| DNN | 0.9737 | 0.4530 | 0.3472 | 0.9731 | 0.4584 | 0.3508 |
| GW-DNN | 0.9885 | 0.2990 | 0.2250 | 0.9871 | 0.3189 | 0.2399 |

**Note.** This table compares the performance metrics of introduced machine learning models and their geographically weighted counterparts

The comparison between the models presents that the OLS model could achieve R² value of 0.8985 and the model is able to explain about 90 percent of the variation in the cancer prevalence rate. However, the values of RMSE (0.8919) and MAE (0.6687) show that this model is not effective in capturing local geographic differences. Unlike OLS, GWR model could reach higher R² and values of RMSE and MAE decreased. This improvement shows that GWR model can represent spatial differences better.

A much powerful predictive capability was observed when ensemble machine learning models were developed. The RF model could perform better than OLS and GWR with R² of 0.9858 for training set and R^²^ of 0.9543 for testing set. Additionally, the values of RMSE (0.5935), and MAE (0.4372) also dropped. These results demonstrated that RF could capture the nonlinear relationships between the variables more effectively. However, the difference between the training and testing results displays some signs of overfitting. Adding spatial weight to the RF model led to better results with R² of 0.9958 for training set and a R² of 0.9661 for testing set, with improvement in RMSE (0.5108) and MAE (0.3777). So, this type of model not only can find nonlinear relationships, but it is able to identify the spatial differences as well.

XGBoost model had advanced predictive performance, and it became as one of the strongest models in this analysis. The R² of 0.9734 for testing set and the values of RMSE (0.4525) and MAE (0.3414) presented high performance of this model in generalizing on unseen data. Furthermore, the geographically weighted version of this model even reached the higher R² value of 0.9971 for the testing set. It also produces lower values for RMSE (0.3902) and MAE (0.2887). So, integrating geographically weighted mechanisms into the boosting model helps to capture the local spatial patterns.

The DNN model exhibited relatively weaker performance, and this model was less effective than RF, and XGBoost. This may be due to the conventional architecture of DNN which require large datasets to be optimized in their learning process. However, when spatial relationships were incorporated into the model, their predictive ability improved dramatically. The GW-DNN achieved a R² of 0.9874 for testing set, and the RMSE and MAE values reduced to 0.3189 and 0.2399, respectively. These findings demonstrated that spatial weighting significantly improves the ability of models by adapting the learning process to local spatial conditions.

### Local goodness of fit

In addition to the aggregated R² values, Figure 5 displays the geographic distribution of local goodness-of-fit (pseudo-R²) for these models. The GW-RF model illustrated improved distribution of pseudo-R² values compared to statistical GWR model. The GW-XGBoost model presented further enhanced performance and larger proportions of the census tracts attained higher values exceeding 97 percent. Although the GW-DNN model achieved a slightly higher overall R² on the testing dataset than GW-XGBoost, the spatial distribution of its local pseudo-R² values tells a different story. The model exhibited inconsistent performance across geographic locations, with some census tracts producing relatively low values, including negative pseudo-R² values in certain areas. In addition, several regions could not reach pseudo-R² values above 0.80. These findings highlight the superior ability of GW-XGBoost compared to all other models.

**Figure 5.**
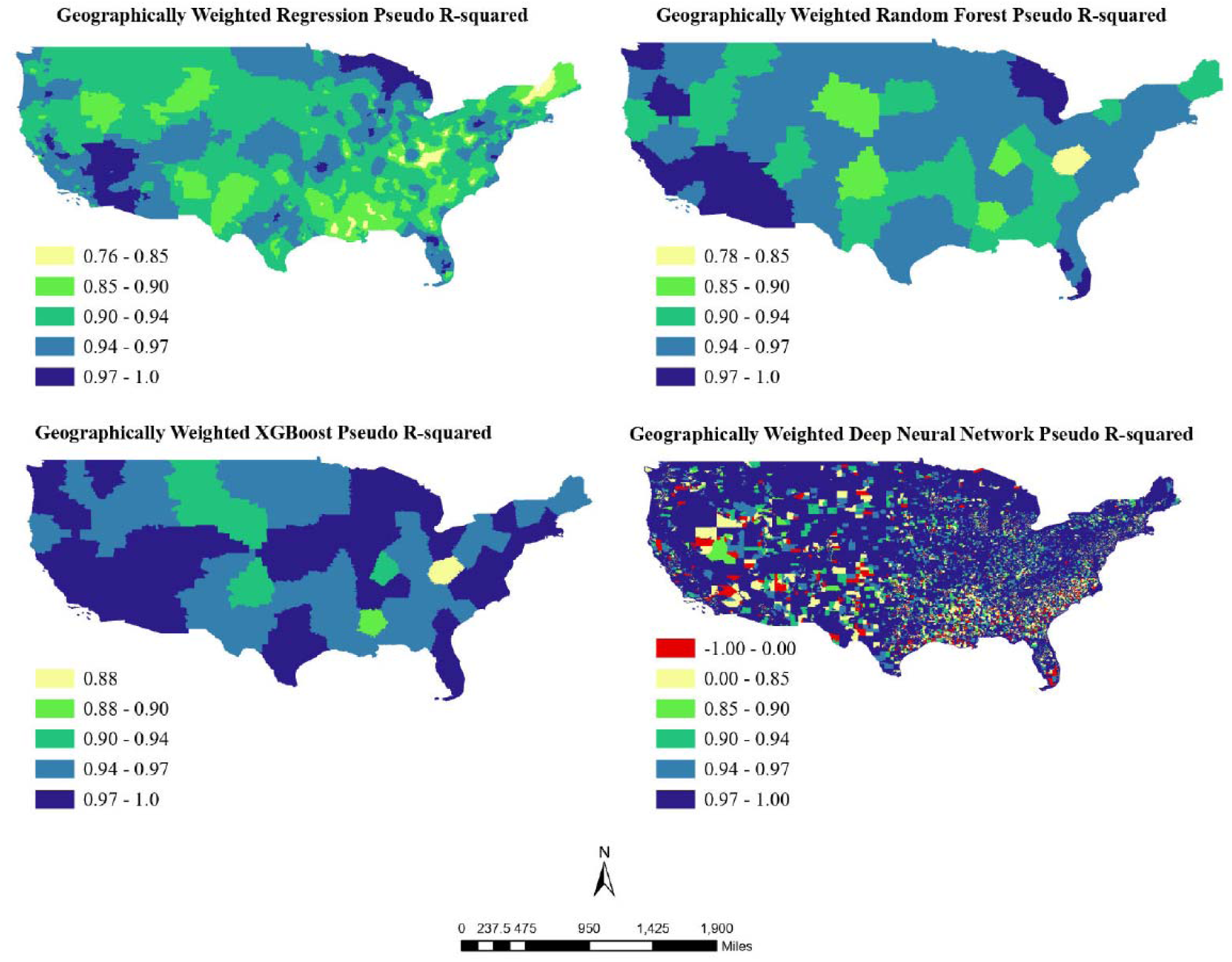
Local Goodness of fit for geographically weighted models at the census tract level

### Feature Importance and Spatial Heterogeneity

Based on the major divisions used for feature importance analysis, we tried to find the mean normalized gain of the top 10 most important features in the whole U.S. (Figure 6). This analysis indicates that percentage of aged people older than 60 years old and percentage of white residents showed the highest normalized gain values among all demographic factors. Besides that, preventative behavioral factors including colorectal cancer screening, and annual medical checkups showed significant impact on the model performance. Metabolic and behavioral determinants such as high cholesterol, diabetes, and hypertension illustrated meaningful contribution to the model performance.

**Figure 6.**
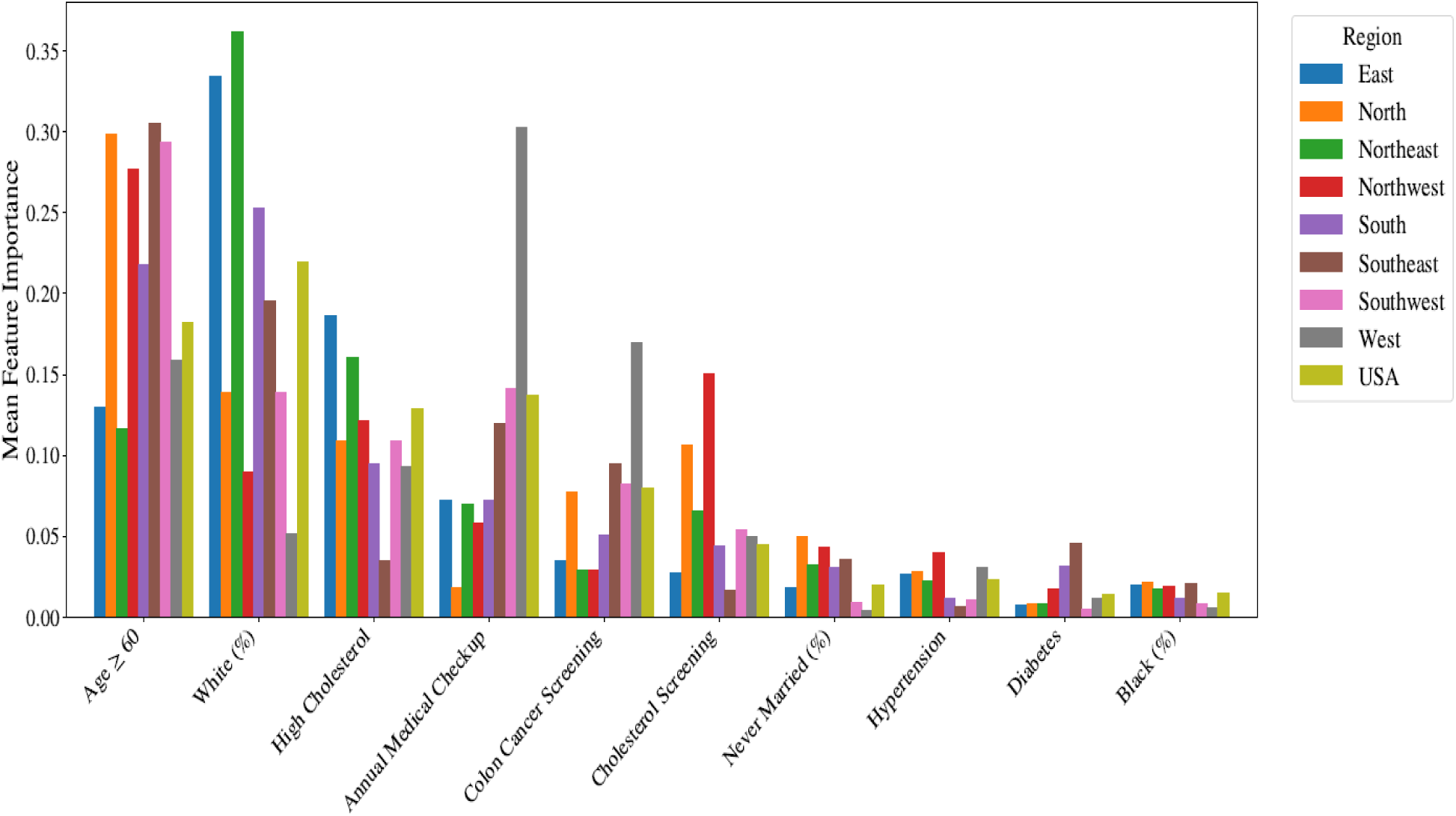
Regional and National Feature Importance Patterns in the GW-XGBoost Model

The analysis of spatial patterns shows that there are significant differences in regional impact of factors that are strongly associated with cancer prevalence. As it is shown in Figure 6 and Figure 7 the relative importance of these factors varied markedly in different parts of the United States. These figures show that the percentage of adult people more than 60 years old is one of the top predictors in all regions, typically ranked between first to third. The influence of other factors is more specific to the region. For example, diabetes was shown as the top five predictors in the Southeast, but it did not appear even among the top ten most influential predictors in other regions such as Northwest and Northeast.

**Figure 7.**
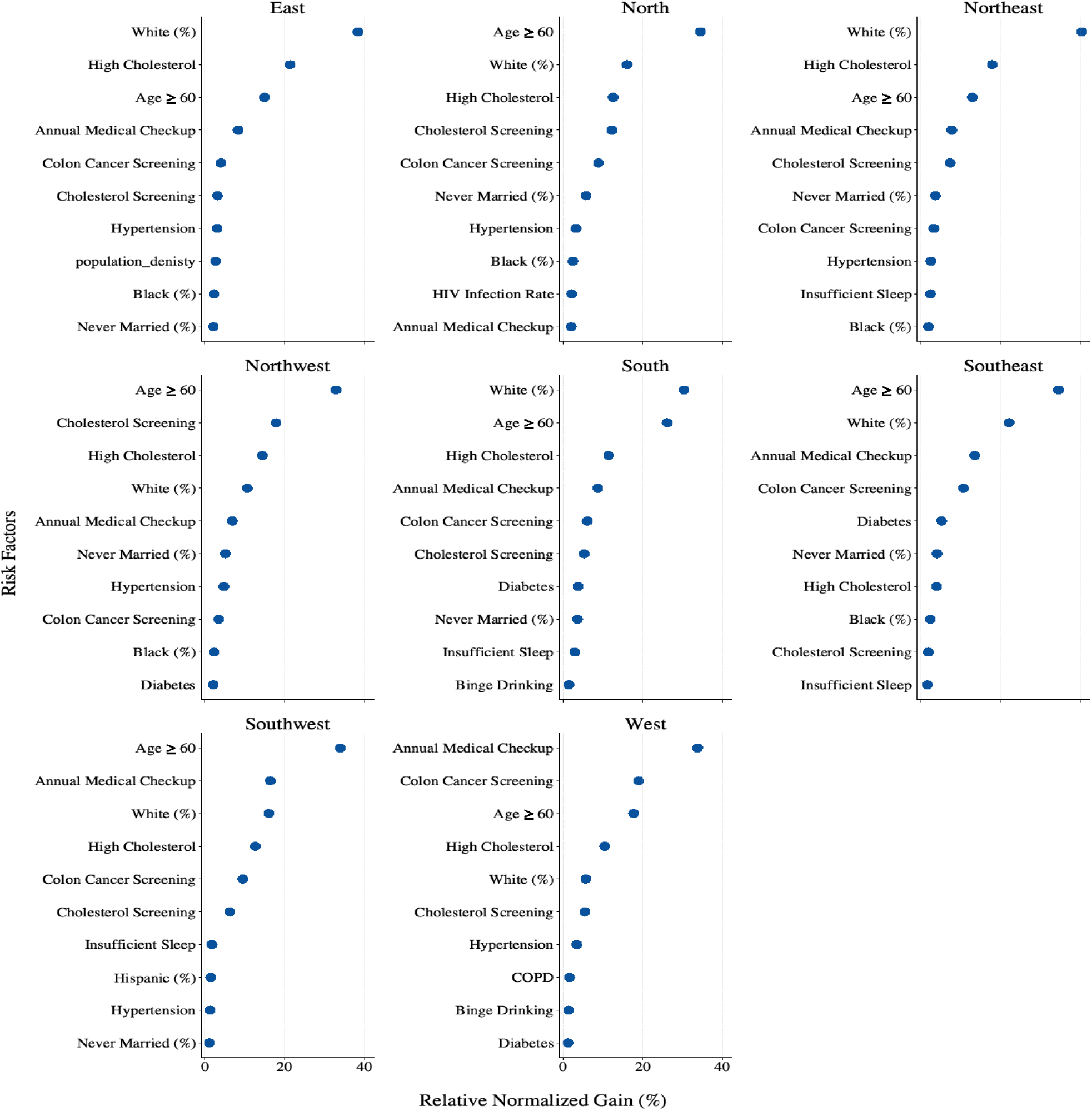
Spatial Heterogeneity in Dominant Risk Factors for Cancer Prevalence Across U.S. Regions. *Note.* Relative normalized gain (%) presents the contribution of each predictor to the trained geographically weighted XGBoost model within each U.S. region. Higher values indicate greater regional importance of the corresponding risk factor.

Notably, the distribution of cancer prevalence across the country is led by complex interactions of multiple risk factors rather than their separate magnitudes. For instance, the percentage of aged people is higher in the Noth, Northeast and parts of the west. However, this factor is highly influential across all regions regardless of its absolute prevalence. Similarly, the prevalence of annual routine medical checkups is higher in the Southeast than in the southwest, but its relative importance is greater in the southwest. A similar pattern is also observed for racial factors. Despite a lower percentage of white individuals in the South compared to the North part of the country, this predictor shows higher relative importance in the Southern region (Figure 7 and Figure 8 and Figure 9). These contradictory patterns are also observable for hypertension in the Southeast and Southwest parts of the United States.

**Figure 8.**
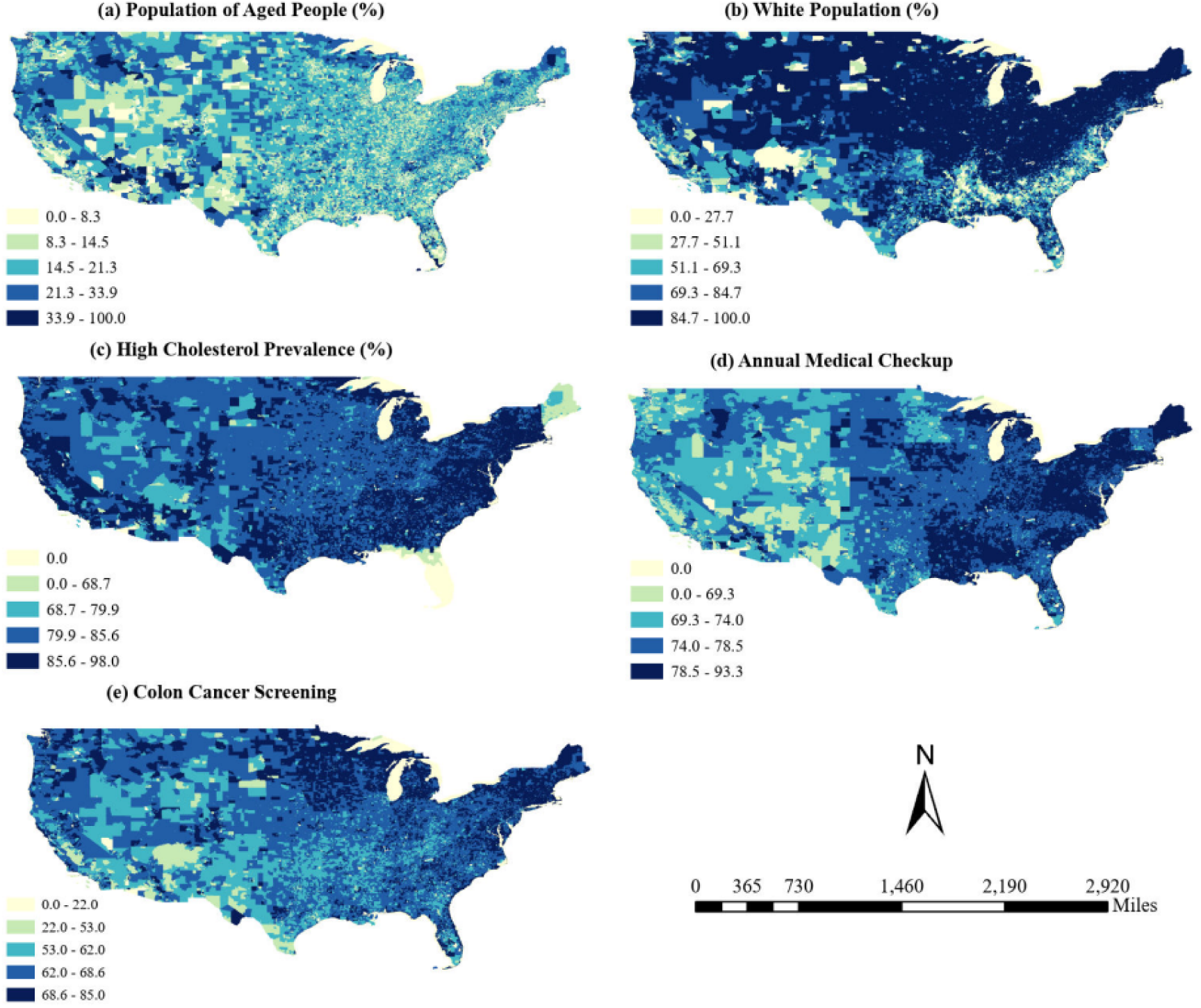
Spatial Distribution of the First Five Most Influential Predictors Identified by the GW-XGBoost Model. *Note.* Geographic distribution of selected demographic and preventive healthcare factors across U.S. census tracts: (a) population aged ≥60 years, (b) percentage of white population, (c) high cholesterol prevalence, (d) annual medical checkup prevalence, and (e) colon cancer screening

**Figure 9.**
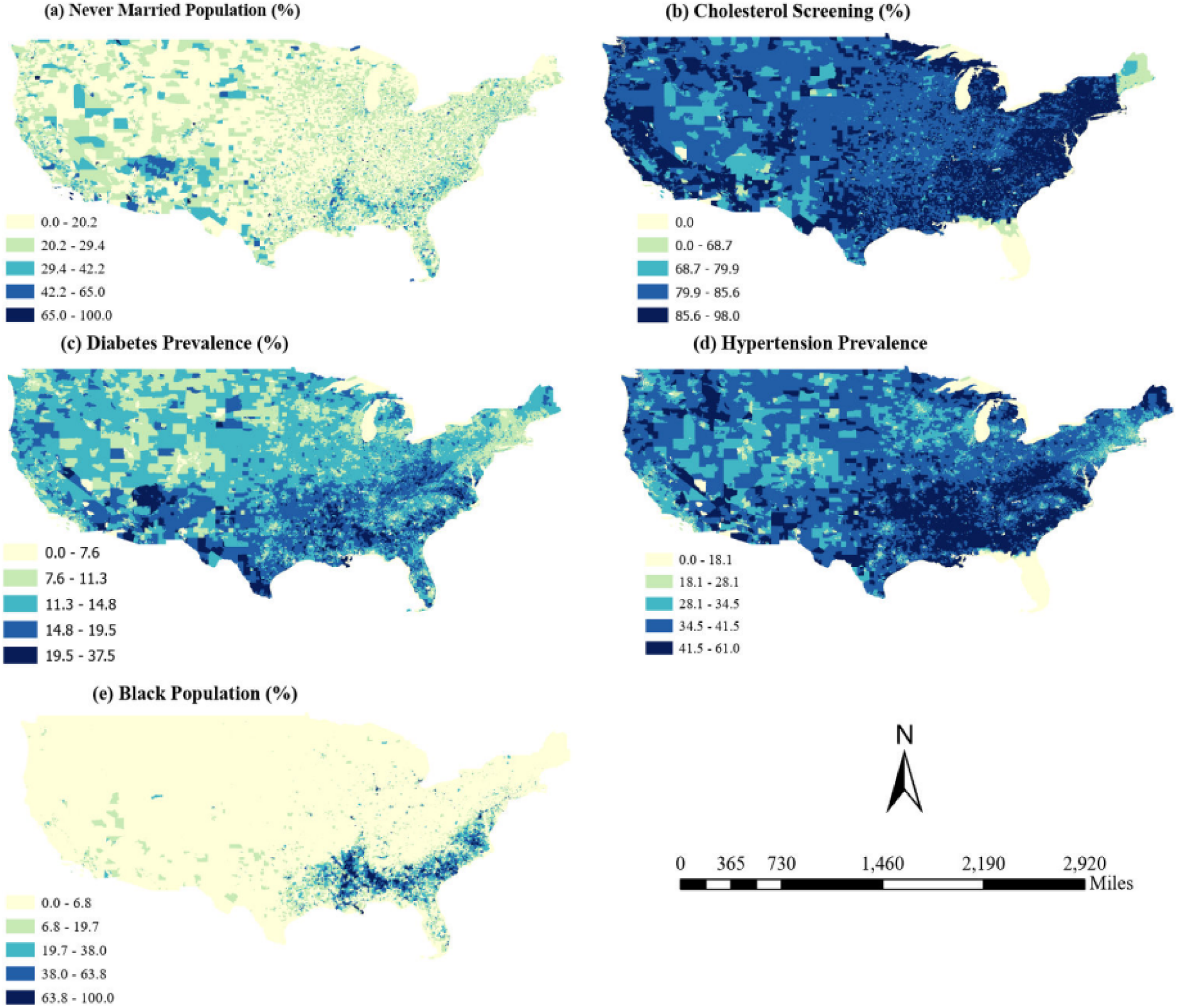
Spatial Distribution of the Second Five Most Influential Predictors Identified by the GW-XGBoost Model. *Note.* Maps illustrate the percentage of (a) never married population, (b) cholesterol screening, (c) diabetes, (d) hypertension, and (e) Black population across U.S. census tracts.

Further analysis of importance plot illustrates the diversity of local cancer drivers at the level of census tract. Figure 10 displays feature importance plots of three separate census tracts in different states including Virginia, Florida, and Texas which were randomly selected from areas with high cancer prevalence rates. These plots show that several major factors such as percentage of aged people and white individuals, besides preventative behavioral factors such as colorectal cancer screening and annual medical checkups are influential in those three tracts.

**Figure 10.**
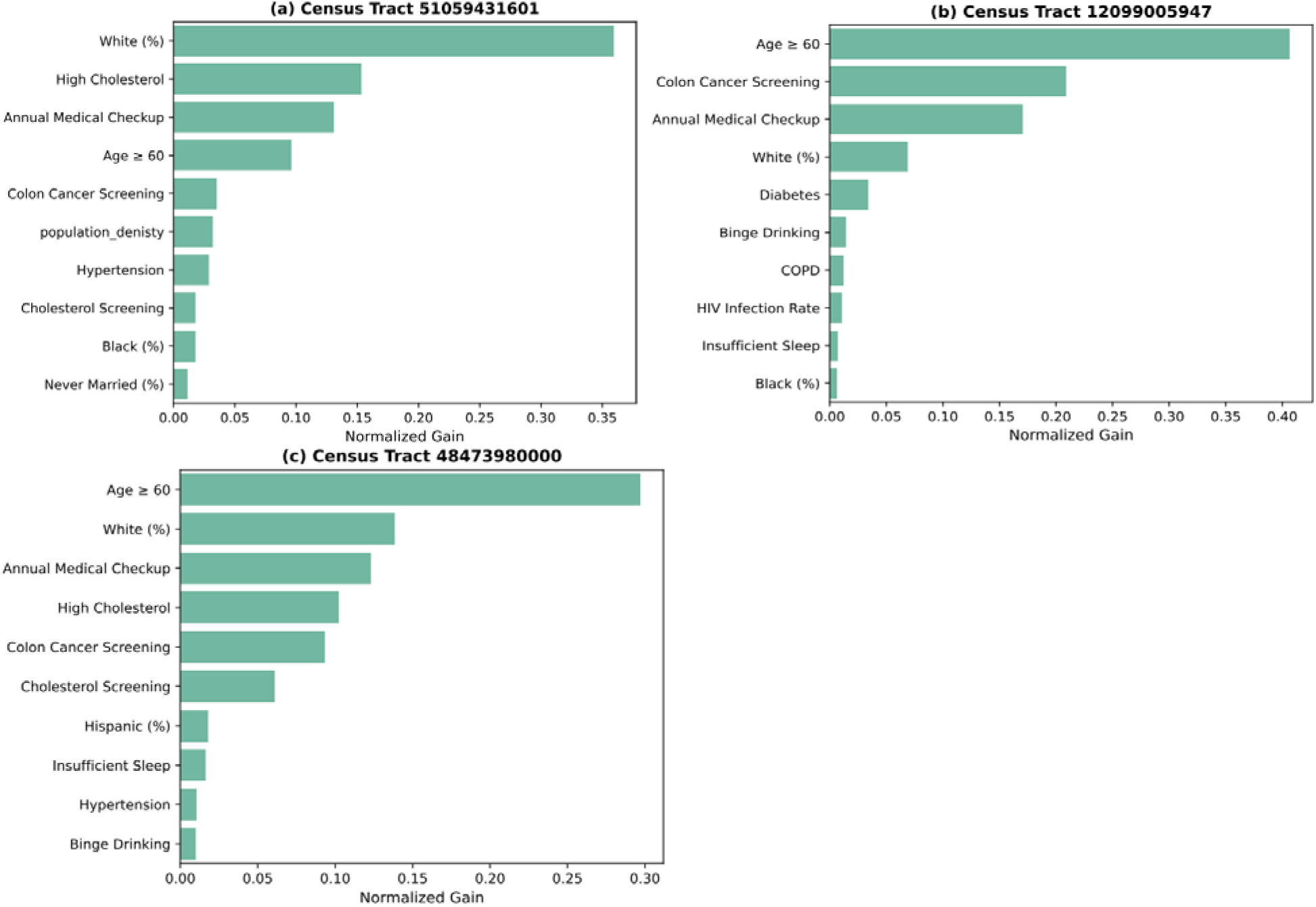
GW-XGBoost feature importance for selected high-prevalence census tracts. *Note.* This figure presents GW-XGBoost feature importance for selected high-prevalence census tracts: a) Fairfax County, Virginia (GEOID: 51059431601), b) Palm Beach County, Florida (GEOID: 12099005947); and c) a census tract in Texas Waller County (GEOID: 48473980000). Bars show normalized gain values, and greater values indicate higher importance of specific predictors in explaining cancer prevalence.

However, the relative importance of these factors significantly changed by location. Some factors, such as COPD prevalence and the proportion of Hispanic residents, were influential only in specific census tracts and did not consistently contribute across all study areas.

## Discussion

This study shows that cancer prevalence is different across census tracts and that the relationships among factors such as population characteristics, healthcare access, lifestyle behaviors, health conditions, and environmental exposures do not follow uniform patterns and can shape this spatial heterogeneity. By applying geographically weighted machine learning models, we show that importance and impact of these cancer risk factors change from one area to another even within the same county.

Compared with previous studies that used machine learning models in the geographical analysis of cancer, this study demonstrated much higher local model performance at the small geographic scale of census tracts. Earlier research by Dong et al. (2022) which used ensemble models at the county level, reported local pseudo-R² values ranging from 0.00 to 0.74. Similarly, Gu et al. (2023) deployed geographically weighted random forest model in China and achieved pseudo-R² values between 0.16 and 0.60 at the provincial level. In another study, Hashtarkhani et al. (2025) used global models at the census tract level to find mammography screening prevalence and reported an R² value of 0.645. However, the GW-XGBoost model used in this study could achieve higher local goodness of fit and the pseudo-R² values of most regions exceeded 0.94 and even reach as high as 0.98 in some groups of census tracts.

Our results also show the increased capability of geographically weighted models in capturing the local spatial differences in cancer prevalence compared to global models. All the geographically weighted models could achieve better performance compared to their global peers. Among all developed predictive models, GW-XGBoost model achieved the highest R² as well as robust and high local R² values which shows its powerful ability to learn complex local spatial patterns. However, the performance metrics showed an important limitation. As presented in Table 3, the RMSE of the test set was approximately twice the training set. This shows signs of overfitting and presence of spatial autocorrelation in the training dataset. So, the model could learn delicate spatial structures, but some of these learned patterns may not generalize well on unseen data.

Also, a methodological difference existed between the geographically weighted ensemble models and GW-DNN that should be considered when interpreting their performance. In the geographically weighted ensemble models, the train set was selected according to geographic proximity and spatial correlation, resulting in overfitting in these types of models. In contrast, the GW-DNN used conventional random training and testing split, and it could learn different spatial relationships. Because of this difference, the comparison between these two approaches should be interpreted with caution.

Another notable finding of this study is that the contribution of factors in cancer prevalence are not the same in all regions, although some factors were influential across most areas. Cancer prevalence was strongly influenced by population characteristics, especially the percentage of older adults and white populations. In addition, preventive healthcare behaviors, such as colorectal cancer screening and regular medical checkups, also played an important role on cancer prevalence at the census tract level. However, behavioral and metabolic risk factors such as diabetes, hypertension, high cholesterol, binge drinking, and insufficient sleep had strong impact on cancer prevalence, but their influence varied substantially by location.

Furthermore, the importance of risk factors does not even consistently align with their prevalent distributions. Some predictors with higher prevalence showed strong association only in particular regions. Other areas showed high sensitivity to factors which were not relatively common locally. This difference presented that feature importance exhibits the local response of cancer prevalence to changes in a factor rather than the absolute prevalence of that factor itself. These results are also aligned by the study conducted by Dong e t al. (2022) at the county level.

Since the impact of risk factors are different in various regions, public health strategies should be fit to the specific demographic, behavioral and metabolic features of each region rather than using a uniform national plan. Increasing population access to regular medical checkups and especially cancer screening programs can improve early detection of cancer. Relevant efforts to increase access to healthcare should focus more on poor communities and older adults who are more vulnerable to this disease. According to the strong association of the percentage of old and white people with cancer prevalence, the educational programs and prevention campaigns should be designed for these high-risk groups.

Additionally, due to the strong association of metabolic diseases such as diabetes, hypertension, and high cholesterol with burden of cancer, it is required that early diagnosis, monitoring, and management of these metabolic conditions will be promoted in the regions with higher impacts. Also, healthier behaviors such as reduction in binge drinking should be encouraged. Improvement in sleep duration and quality, as well as increase of physical activity, and adopting healthier diets need community-based interventions that can lower the risk of cancer. Educational campaigns can also increase public awareness about modifiable cancer risk factors and emphasize the importance of routine screening, having a healthy lifestyle, and use of preventative healthcare. Policy makers need to allocate healthcare resources and prevention programs to areas with high levels of unhealthy behaviors or metabolic health problems. Our findings highlighted that spatial and neighborhood level analysis can guide the public health planning to the correct direction and interventions can more effectively address the local high burden of cancer and its risk factors.

## Conclusion

This study showed that geographically weighted machine learning models can identify the localized spatial patterns more effectively than global learners in cancer prevalence and its determinants especially at smaller geographic levels. Among all the developed machine learning models, GW-XGBoost had the highest performance with some specific limitations. This approach presented that major risk factors of cancer varied location by location and these differences require resilient public health system to allocate resources according to essential needs of each specific region or community.

Despite meaningful results obtained in this study, this analysis relies on cross-sectional data and did not deploy longitudinal information. Therefore, temporal changes or dynamic interactions of risk factors over time haven’t been considered. Furthermore, data of cancer prevalence and some behavioral, health conditions, and metabolic factors were derived from the CDC PLACES dataset. This dataset provides model-based estimates rather than direct surveillance, and this increases uncertainty. Also, we used an ecological design that performed at the population level. So, results are subject to ecological fallacy and therefore cannot be used to infer the risk of cancer at the individual level. Additionally, several variables examined in this study were measured at a broad level and did not distinguish between specific subtypes or conditions, such as different forms of cancer, diabetes, hypertension, and other health-related factors.

Based on the limitations of this study including the lack of reliable observed datasets at this geographic scale, the absence of longitudinal analysis, and use of population level data, future research can incorporate longitudinal datasets and validate model predictions using individual level clinical data where feasible. Besides that, the use of higher quality observed datasets can improve the generalizability of the results to real world settings. So, using geographically weighted machine learning models to longitudinal data can enhance the predictive performance of these models and can help deeper understanding of major cancer risk factors affecting cancer prevalence overtime.

## Data Availability

All data produced in the present study are available upon reasonable request to the authors

## Declaration of generative AI use

In this manuscript, generative AI was used for grammar checking, improving readability and improving Python code development.

## Appendix A Summary of the Studied Variables and Data Sources

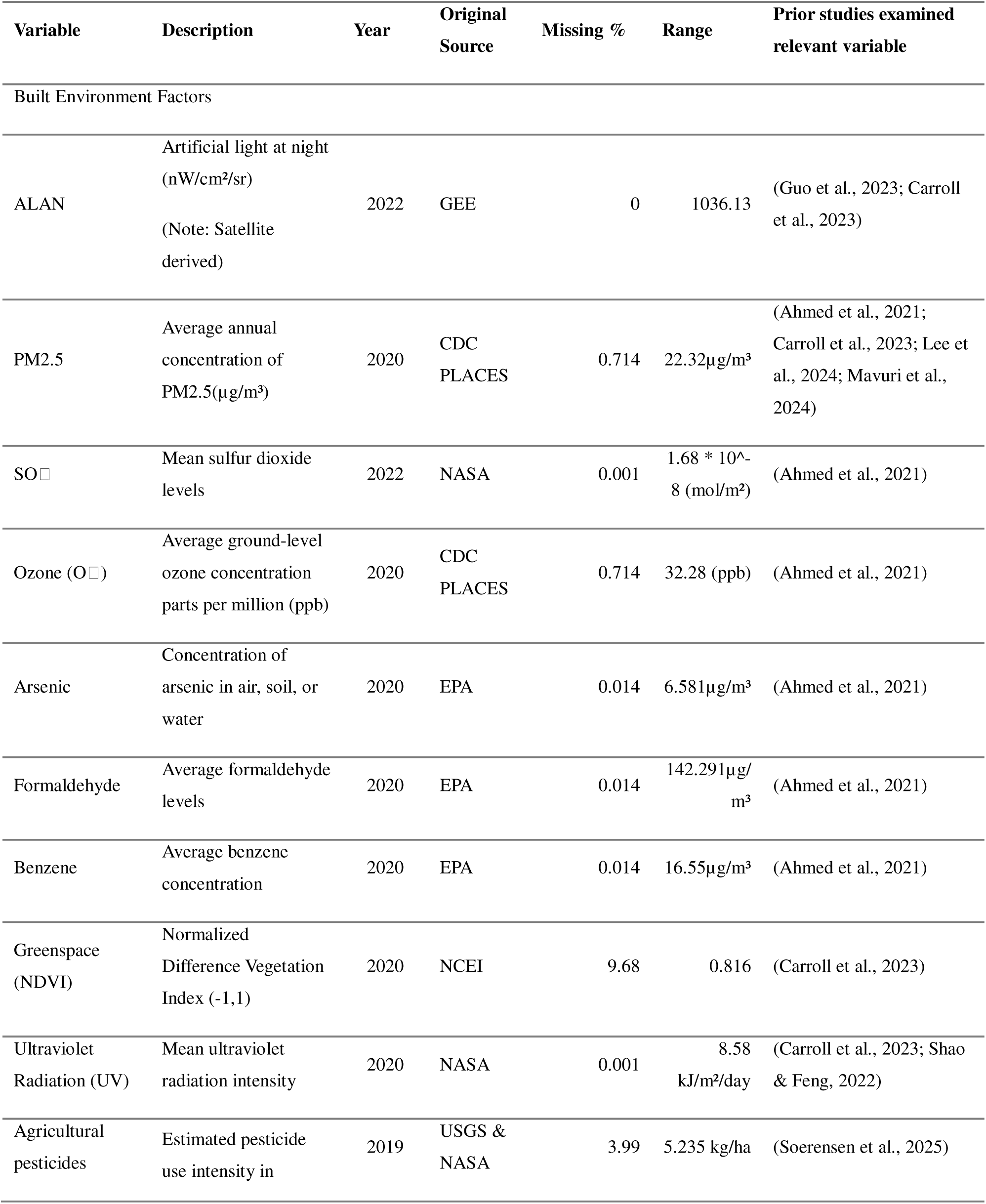
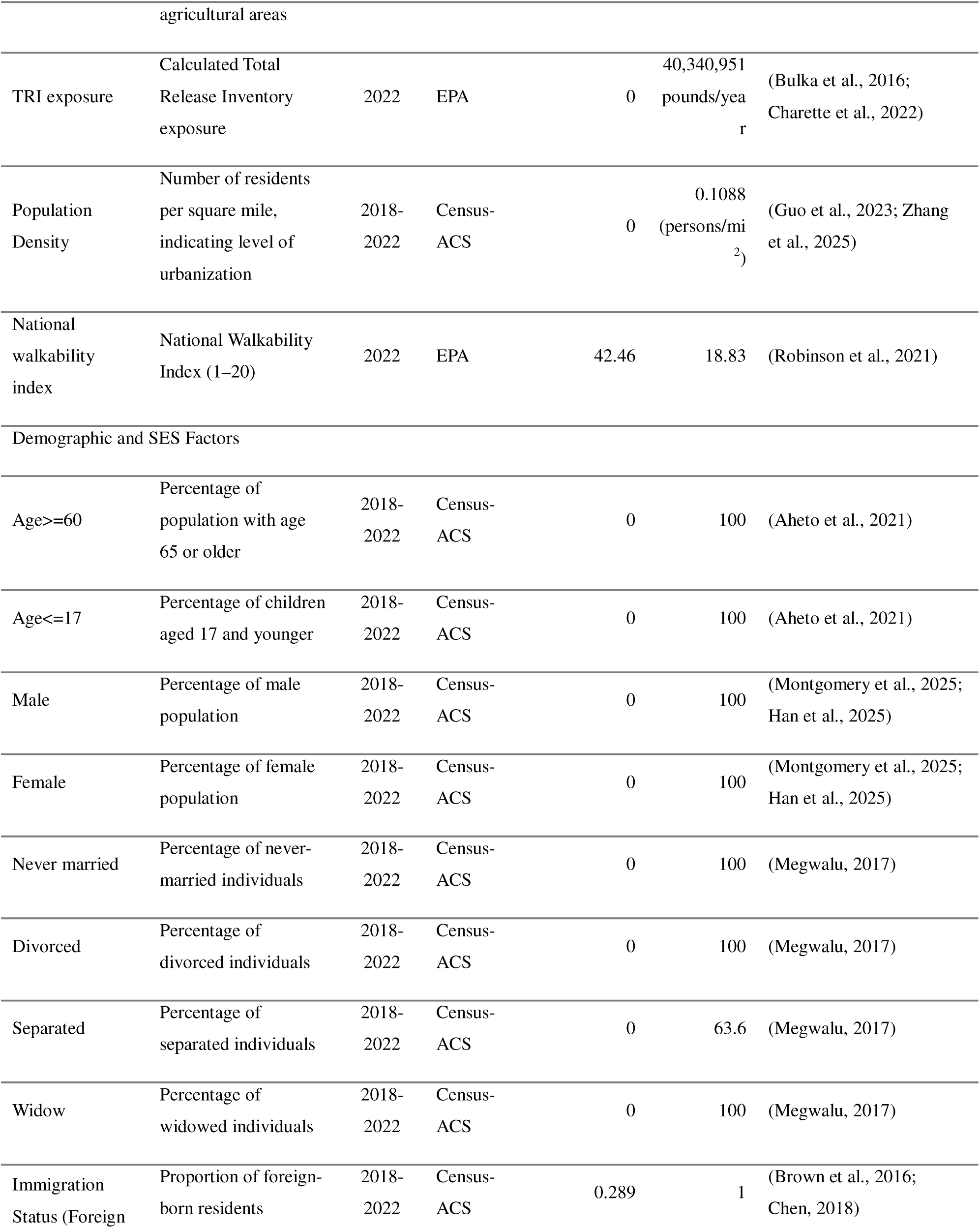
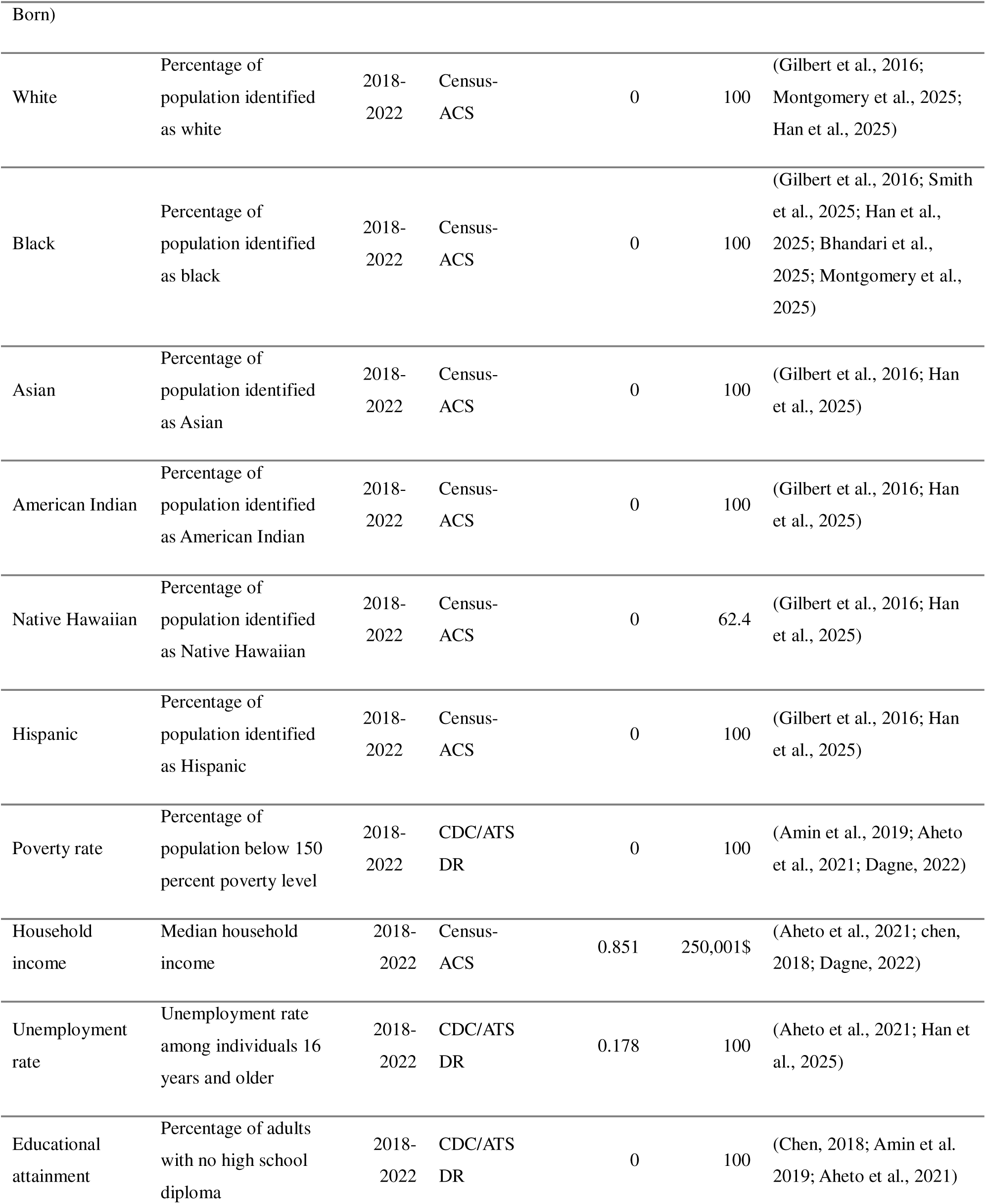
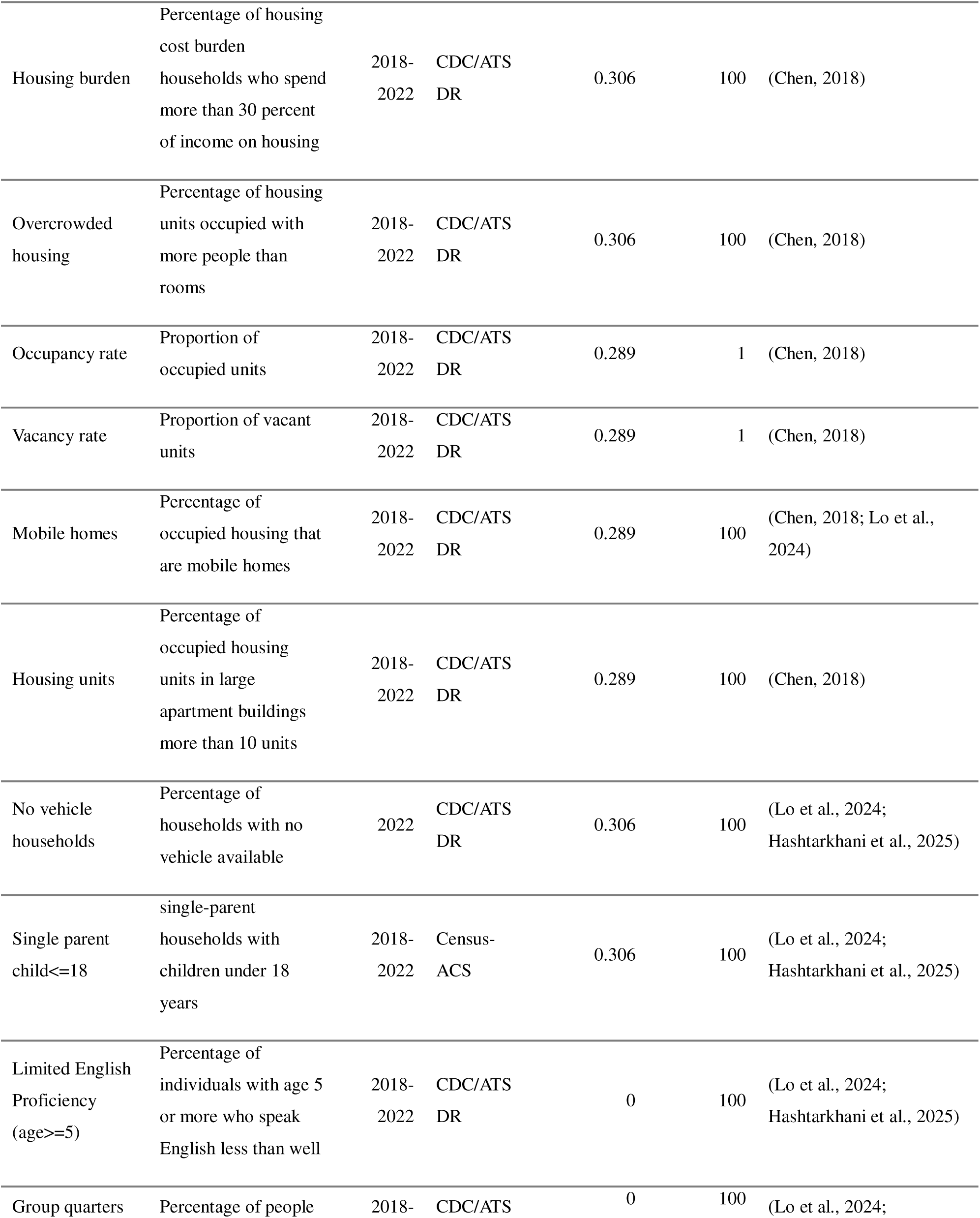
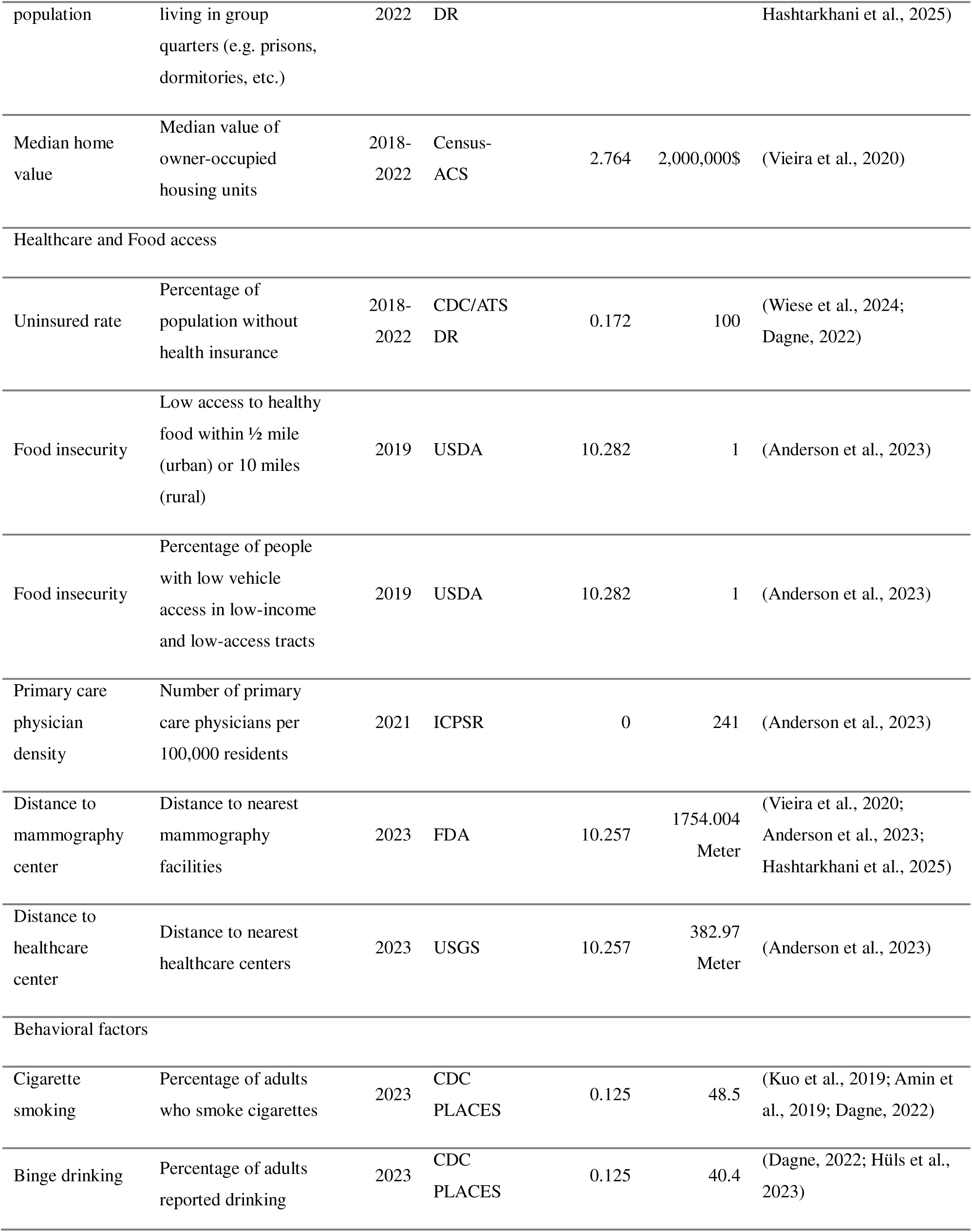
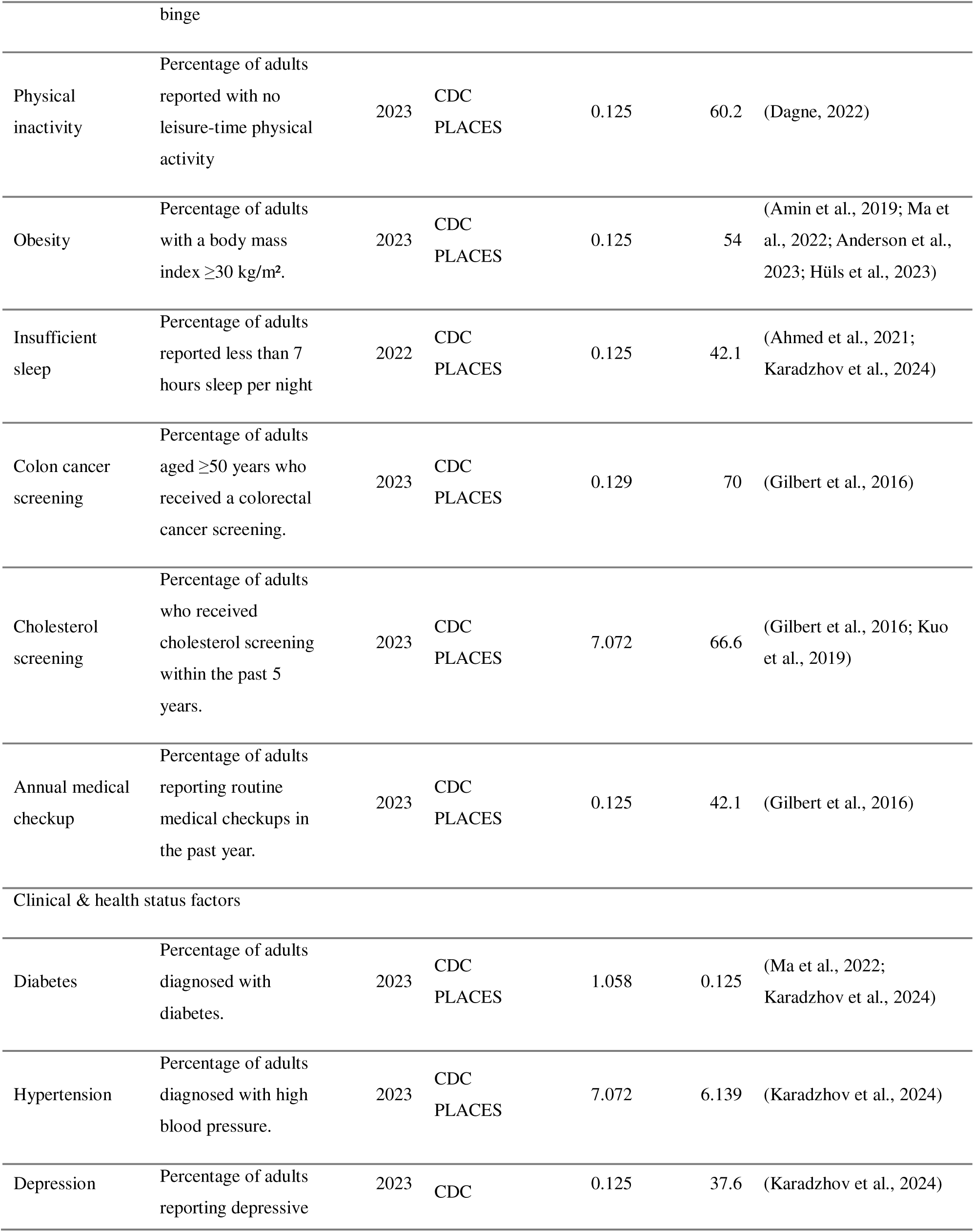
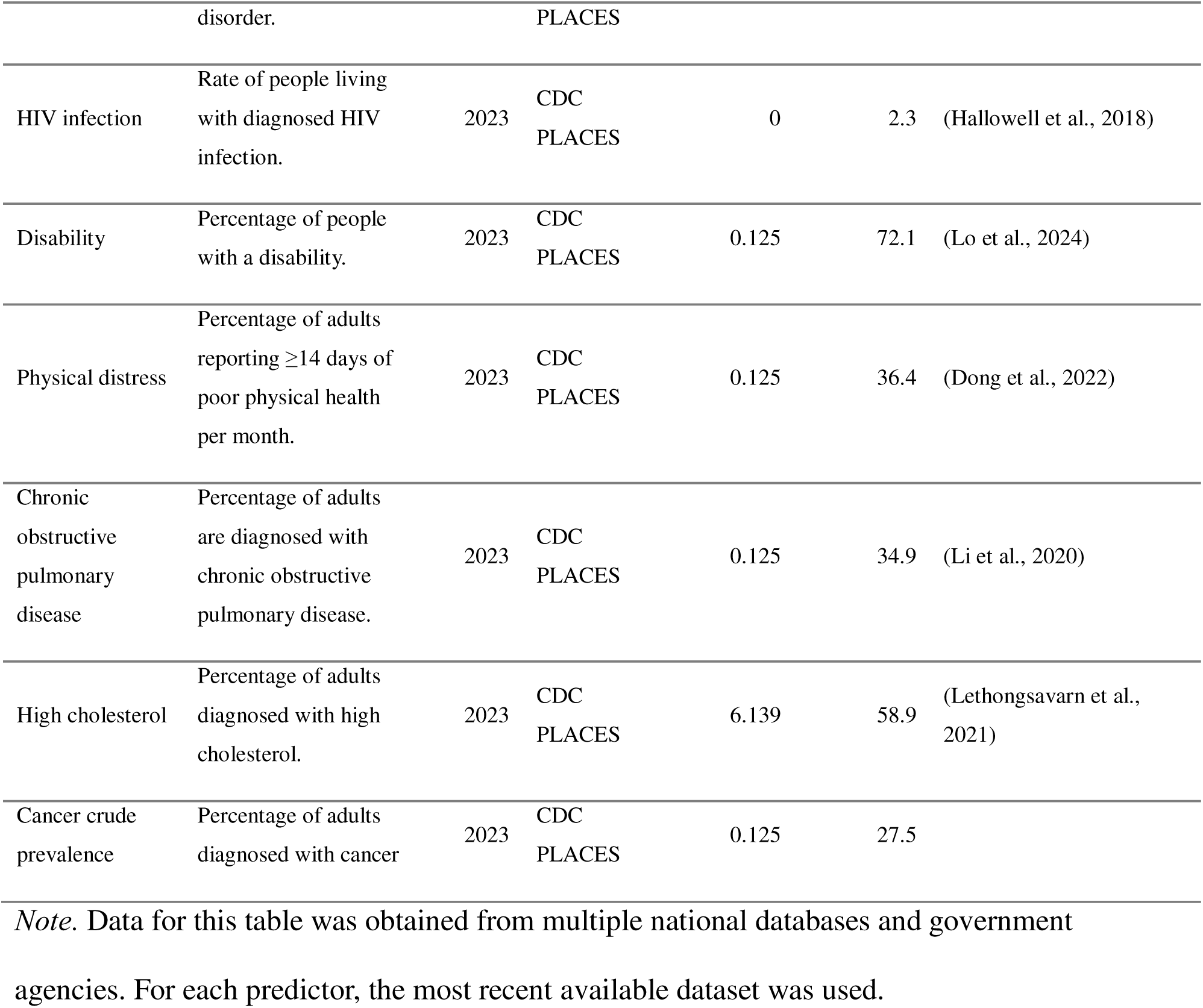

## Appendix B Eight major Divisions of the United States

| State | East | North | Northeast | Northwest | South | Southeast | Southwest | West |
| --- | --- | --- | --- | --- | --- | --- | --- | --- |
| Alabama | 1 | 0 | 0 | 0 | 0 | 1436 | 0 | 0 |
| Alaska | 1 | 0 | 0 | 82 | 0 | 0 | 0 | 94 |
| Arizona | 0 | 0 | 0 | 0 | 0 | 0 | 0 | 1765 |
| Arkansas | 0 | 0 | 0 | 0 | 418 | 108 | 295 | 2 |
| California | 0 | 0 | 0 | 0 | 0 | 0 | 0 | 9129 |
| Colorado | 0 | 0 | 0 | 0 | 0 | 0 | 0 | 1447 |
| Connecticut | 884 | 0 | 0 | 0 | 0 | 0 | 0 | 0 |
| Delaware | 262 | 0 | 0 | 0 | 0 | 0 | 0 | 0 |
| District of<br>Columbia | 206 | 0 | 0 | 0 | 0 | 0 | 0 | 0 |
| Florida | 0 | 0 | 0 | 0 | 0 | 5160 | 0 | 0 |
| Georgia | 312 | 0 | 0 | 0 | 0 | 2484 | 0 | 0 |
| Hawaii | 0 | 0 | 0 | 0 | 0 | 0 | 0 | 461 |
| Idaho | 0 | 0 | 0 | 25 | 0 | 0 | 0 | 431 |
| Illinois | 167 | 105 | 2993 | 0 | 0 | 0 | 0 | 0 |
| Indiana | 727 | 0 | 969 | 0 | 0 | 0 | 0 | 0 |
| Iowa | 0 | 540 | 0 | 356 | 0 | 0 | 0 | 0 |
| Kansas | 0 | 0 | 0 | 303 | 0 | 0 | 0 | 526 |
| Kentucky | 1306 | 0 | 0 | 0 | 0 | 0 | 0 | 0 |
| Louisiana | 0 | 0 | 0 | 0 | 1381 | 0 | 7 | 0 |
| Maine | 404 | 0 | 3 | 0 | 0 | 0 | 0 | 0 |
| Maryland | 1475 | 0 | 0 | 0 | 0 | 0 | 0 | 0 |
| Massachusetts | 1620 | 0 | 0 | 0 | 0 | 0 | 0 | 0 |
| Michigan | 0 | 33 | 2984 | 0 | 0 | 0 | 0 | 0 |
| Minnesota | 0 | 1351 | 0 | 154 | 0 | 0 | 0 | 0 |
| Mississippi | 0 | 0 | 0 | 0 | 419 | 459 | 0 | 0 |
| Missouri | 96 | 152 | 544 | 541 | 13 | 42 | 46 | 220 |
| Montana | 0 | 0 | 0 | 281 | 0 | 0 | 0 | 38 |
| Nebraska | 0 | 0 | 0 | 504 | 0 | 0 | 0 | 49 |
| Nevada | 0 | 0 | 0 | 0 | 0 | 0 | 0 | 779 |
| New<br>Hampshire | 350 | 0 | 0 | 0 | 0 | 0 | 0 | 0 |
| New Jersey | 2181 | 0 | 0 | 0 | 0 | 0 | 0 | 0 |
| New Mexico | 0 | 0 | 0 | 0 | 0 | 0 | 15 | 597 |
| New York | 5138 | 0 | 273 | 0 | 0 | 0 | 0 | 0 |
| North Carolina | 2672 | 0 | 0 | 0 | 0 | 0 | 0 | 0 |
| North Dakota | 0 | 0 | 0 | 228 | 0 | 0 | 0 | 0 |
| Ohio | 2798 | 0 | 370 | 0 | 0 | 0 | 0 | 0 |
| Oklahoma | 0 | 0 | 0 | 0 | 0 | 0 | 309 | 896 |
| Oregon | 0 | 0 | 0 | 0 | 0 | 0 | 0 | 1001 |
| Pennsylvania | 3446 | 0 | 0 | 0 | 0 | 0 | 0 | 0 |
| Rhode Island | 250 | 0 | 0 | 0 | 0 | 0 | 0 | 0 |
| South Carolina | 1253 | 0 | 0 | 0 | 0 | 70 | 0 | 0 |
| South Dakota | 0 | 0 | 0 | 242 | 0 | 0 | 0 | 0 |
| Tennessee | 1257 | 0 | 0 | 0 | 0 | 444 | 0 | 0 |
| Texas | 0 | 0 | 0 | 0 | 332 | 0 | 6114 | 450 |
| Utah | 0 | 0 | 0 | 0 | 0 | 0 | 0 | 716 |
| Vermont | 193 | 0 | 0 | 0 | 0 | 0 | 0 | 0 |
| Virginia | 2198 | 0 | 0 | 0 | 0 | 0 | 0 | 0 |
| Washington | 0 | 0 | 0 | 17 | 0 | 0 | 0 | 1767 |
| West Virginia | 546 | 0 | 0 | 0 | 0 | 0 | 0 | 0 |
| Wisconsin | 0 | 460 | 1082 | 0 | 0 | 0 | 0 | 0 |
| Wyoming | 0 | 0 | 0 | 30 | 0 | 0 | 0 | 130 |

*Note.* These eight regions were defined based on each tract's direction related to the geographic center of the United States and calculated from the mean latitude and longitude of all tract centroids. Census tracts were assigned to one of eight 45° sectors using their angular position.

## References

Aheto, J. M. K., Utuama, O. A., & Dagne, G. A. (2021). Geospatial analysis, web-based mapping and determinants of prostate cancer incidence in Georgia counties: Evidence from the 2012–2016 SEER data. BMC Cancer, 21(1), 508. 10.1186/s12885-021-08254-0

Ahmed, Z. U., Sun, K., Shelly, M., & Mu, L. (2021). Explainable artificial intelligence (XAI) for exploring spatial variability of lung and bronchus cancer (LBC) mortality rates in the contiguous USA. Scientific Reports, 11(1), 24090. 10.1038/s41598-021-03198-8

Alsahaf, A., Petkov, N., Shenoy, V., & Azzopardi, G. (2022). A framework for feature selection through boosting. Expert Systems with Applications, 187, 115895. 10.1016/j.eswa.2021.115895

Amin, R. W., Ross, A. M., Lee, J., Guy, J., & Stafford, B. (2019). Patterns of ovarian cancer and uterine cancer mortality and incidence in the contiguous USA. Science of the Total Environment, 697, 134128. 10.1016/j.scitotenv.2019.134128

Anderson, T., Herrera, D., Mireku, F., Barner, K., Kokkinakis, A., Dao, H., Webber, A., Diaz Merida, A., Gallo, T., & Pierobon, M. (2023). Geographical variation in social determinants of female breast cancer mortality across US counties. JAMA Network Open, 6(9), e2333618–e2333618. 10.1001/jamanetworkopen.2023.33618

Bhandari, Anish; Deng, Shawn; Olheiser, Michael; and Slater, Robert (2025) Predictive modeling of colorectal cancer risk: Leveraging health, demographic, and socioeconomic factors for targeted screening, SMU Data Science Review, 9(1). https://scholar.smu.edu/datasciencereview/vol9/iss1/5

Bhattacharya, M., Cronin, K. A., Farrigan, T. L., Kennedy, A. E., Yu, M., & Srinivasan, S. (2024). Description of census-tract–level social determinants of health in cancer surveillance data. JNCI Monographs, 2024(65), 152–161. 10.1093/jncimonographs/lgae027

Bray, F., & Moller, B. (2006). Predicting the future burden of cancer. Nature Reviews Cancer, 6(1), 63–74. 10.1038/nrc1781

Brown, P., Jiang, H., Ezzat, S., & Sawka, A. M. (2016). A detailed spatial analysis on contrasting cancer incidence patterns in thyroid and lung cancer in Toronto women. BMC Public Health, 16(1), 950. 10.1186/s12889-016-3634-4

Buck, K. D. (2016). Modelling of geographic cancer risk factor disparities in US counties. Applied Geography, 75, 28–35. 10.1016/j.apgeog.2016.08.001

Bulka, C., Nastoupil, L. J., Koff, J. L., Bernal-Mizrachi, L., Ward, K., Williams, J. N., Bayakly, A. R., Switchenko, J. M., Waller, L. A., & Flowers, C. R. (2016). Relations between residential proximity to EPA-designated toxic release sites and diffuse large B-cell lymphoma incidence. Southern Medical Journal, 109(10), 606–614. 10.14423/SMJ.0000000000000545

Carroll, R., Ish, J. L., Sandler, D. P., White, A. J., & Zhao, S. (2023). Understanding the role of environmental and socioeconomic factors in the geographic variation of breast cancer risk in the US-wide Sister Study. Environmental Research, 239, 117349. 10.1016/j.envres.2023.117349

Centers for Disease Control and Prevention. (2025). United States cancer statistics: Data visualization. https://gis.cdc.gov/Cancer/USCS/

Charette, A. T., Hill, D. T., Collins, M. B., & Mirowsky, J. E. (2022). Assessing the quantity and toxicity of chemical releases from TRI facilities in Upstate New York. Journal Of Environmental Studies and Sciences, 12(3), 417–429. 10.1007/s13412-022-00759-9

Chen, X. (2018). A spatial and temporal analysis of the socioeconomic factors associated with breast cancer in Illinois using geographically weighted generalized linear regression. Journal of Geovisualization and Spatial Analysis, 2(1), 5. 10.1007/s41651-017-0011-5

Dagne, G. A. (2022). Geographic variation and association of risk factors with incidence of colorectal cancer at small-area level. Cancer Causes & Control, 33(9), 1155–1160. 10.1007/s10552-022-01607-5

Dolatshah, M., Hadian, A., & Minaei-Bidgoli, B. (2015). Ball*-tree: Efficient spatial indexing for constrained nearest-neighbor search in metric spaces. arXiv preprint arXiv:1511.00628. https://arxiv.org/abs/1511.00628

Dong, W., Bensken, W. P., Kim, U., Rose, J., Fan, Q., Schiltz, N. K., Berger, N. A., & Koroukian, S. M. (2022). Variation in and factors associated with US county-level cancer mortality, 2008-2019. JAMA Network Open, 5(9), e2230925–e2230925. 10.1001/jamanetworkopen.2022.30925

Fletcher, S. A., Marchese, M., Cole, A. P., Mahal, B. A., Friedlander, D. F., Krimphove, M., Kilbridge, K. L., Lipsitz, S. R., Nguyen, P. L., Choueiri, T. K., & Kibel, A. S. (2020). Geographic distribution of racial differences in prostate cancer mortality. JAMA Network Open, 3(3), e201839. 10.1001/jamanetworkopen.2020.1839

Gallicchio, L., Devasia, T. P., Tonorezos, E., Mollica, M. A., & Mariotto, A. (2022). Estimation of the number of individuals living with metastatic cancer in the United States. JNCI: Journal of the National Cancer Institute, 114(11), 1476–1483. 10.1093/jnci/djac158

Gilbert, S. M., Pow-Sang, J. M., & Xiao, H. (2016). Geographical factors associated with health disparities in prostate cancer. Cancer Control, 23(4), 401–408. 10.1177/107327481602300411

Gu, W., Xue, F., Han, W., Wang, Z., Zhao, J., Zhang, L., Yang, C., & Jiang, J. (2023). Assessment of the spatial association between multiple pollutants of surface water and digestive cancer incidence in China: A novel application of spatial machine learning. Ecological Indicators, 154, 110897. 10.1016/j.ecolind.2023.110897

Guo, B., Gao, Q., Pei, L., Guo, T., Wang, Y., Wu, H., Zhang, W., & Chen, M. (2023). Exploring the association of PM2. 5 with lung cancer incidence under different climate zones and socioeconomic conditions from 2006 to 2016 in China. Environmental Science and Pollution Research, 30(60), 126165–126177. 10.1007/s11356-023-31138-8

Han, C., Burd, C., Plascak, J., Tounkara, F., Rosko, A., Noonan, A., Tan, A., Von Ah, D., & Ning, X. (2025). Machine learning-based chemotoxicity predictions in patients with colorectal cancer: integrating race, geospatial social determinants of health, and biological aging. Research Square. 10.21203/rs.3.rs-6628340/v1

Hallisey, E., Tai, E., Berens, A., Wilt, G., Peipins, L., Lewis, B., Graham, S., Flanagan, B., & Buchanan Lunsford, N. (2017). Transforming geographic scale: A comparison of combined population and areal weighting to other interpolation methods. International Journal of Health Geographics, 16(1), 29. 10.1186/s12942-017-0102-z

Hallowell, B. D., Robb, S. W., & Kintziger, K. W. (2018). Comparing the geographic distribution and location characteristics of HIV-seropositive and HIV-seronegative individuals with a diagnosis of cancer living in the southeast US. Spatial And Spatio-temporal Epidemiology, 24, 11–18. 10.1016/j.sste.2017.10.002

Hashtarkhani, S., Zhou, Y., Kumsa, F. A., White-Means, S., Schwartz, D. L., & Shaban-Nejad, A. (2025). Analyzing geospatial and socioeconomic disparities in breast Cancer screening among populations in the United States: Machine learning approach. JMIR Cancer, 11, e59882. 10.2196/59882

Hüls, A., Van Cor, S., Christensen, G. M., Li, Z., Liu, Y., Shi, L., Pearce, J. L., Bayakly, R., Lash, T. L., Ward, K., & Switchenko, J. M. (2023). Environmental, social and behavioral risk factors in association with spatial clustering of childhood cancer incidence. Spatial and Spatio-temporal Epidemiology, 45, 100582. 10.1016/j.sste.2023.100582

Islami, F., Guerra, C. E., Minihan, A., Yabroff, K. R., Fedewa, S. A., Sloan, K., Wiedt, T. L., Thomson, B., Siegel, R. L., Nargis, N., Winn, R. A., Lacasse, L., Makaroff, L., Daniels, E. C., Patel, A. V., Cance, W. G., & Jemal, A. (2022). American Cancer Society’s report on the status of cancer disparities in the United States, 2021. CA: A Cancer Journal for Clinicians, 72(2), 112–143. 10.3322/caac.21703

Islam, M. M., Ashik, A. A. M., Islam, S., & Hasan, S. (2025). Geo-spatial analysis of cancer cluster and environmental risk factor in the USA. World Journal of Biomedical Sciences, 3(1), 8–16. 10.61784/wjbs3015

Karadzhov, G., Albert, P. S., Henry, K. A., Abnet, C. C., Lawrence, W. R., Shiels, M. S., Zhang, T., Powell-Wiley, T. M., & Chen, Y. (2024). Cancer mortality and geographic inequalities: A detailed descriptive and spatial analysis of social determinants across US counties, 2018–2021. Public Health, 237, 1–6. 10.1016/j.puhe.2024.08.021

Kuo, T. M., Meyer, A. M., Baggett, C. D., & Olshan, A. F. (2019). Examining determinants of geographic variation in colorectal cancer mortality in North Carolina: A spatial analysis approach. Cancer Epidemiology, 59, 8–14. 10.1016/j.canep.2019.01.002

Lee, H., Hanson, H. A., Logan, J., Maguire, D., Kapadia, A., Dewji, S., & Agasthya, G. (2024). Evaluating county-level lung cancer incidence from environmental radiation exposure, PM2.5, and other exposures with regression and machine learning models. Environmental Geochemistry and Health, 46(3), 82. 10.1007/s10653-023-01820-4

Lethongsavarn, V., Pinault, M., Diedhiou, A., Guimaraes, C., Guibon, R., Bruyère, F., Mathieu, R., Rioux-Leclercq, N., Multigner, L., Brureau, L., Fournier, G., Doucet, L., Blanchet, P., & Fromont, G. (2021). Tissue cholesterol metabolism and prostate cancer aggressiveness: Ethno geographic variations. The Prostate, 81(16), 1365–1373. 10.1002/pros.24234

Li, C., Chen, L., Chou, C., Ngorsuraches, S., & Qian, J. (2022). Using machine learning approaches to predict short-term risk of cardiotoxicity among patients with colorectal cancer after starting fluoropyrimidine-based chemotherapy. Cardiovascular Toxicology, 22(2), 130–140. 10.1007/s12012-021-09708-4

Li, X., Xiao, J., Huang, M., Liu, T., Guo, L., Zeng, W., Chen, Q., Zhang Jim, J., & Ma, W. (2020). Associations of county-level cumulative environmental quality with mortality of chronic obstructive pulmonary disease and mortality of tracheal, bronchus and lung cancers. Science of The Total Environment, 703, 135523. 10.1016/j.scitotenv.2019.135523

Lo, C. H., Tun, K. M., Pan, C. W., Lee, J. K., Singh, H., & Samadder, N. J. (2024). Association between social vulnerability and gastrointestinal cancer mortality in the United States counties. Gastro Hep Advances, 3(6), 821–829. 10.1016/j.gastha.2024.05.007

Luberice, K., Cantrell, M. C., Edgar, L., Smotherman, C., Guerrier, C., Salloum, R. G., Parker, A. S., Mobley, E. M., & Awad, Z. T. (2025). Association of county-level social Determinants and pancreatic cancer incidence in the United States. Anticancer Research, 45(6), 2507–2514. 10.21873/anticanres.17622

Lynch, S. M., Wiese, D., Ortiz, A., Sorice, K. A., Nguyen, M., González, E. T., & Henry, K. A. (2020). Towards precision public health: Geospatial analytics and sensitivity/specificity assessments to inform liver cancer prevention. SSM-Population Health, 12, 100640. 10.1016/j.ssmph.2020.100640

Ma, C., Congly, S. E., Chyou, D. E., Ross-Driscoll, K., Forbes, N., Tsang, E. S., Sussman, D. A., & Goldberg, D. S. (2022). Factors associated with geographic disparities in gastrointestinal cancer mortality in the United States. Gastroenterology, 163(2), 437–448. 10.1053/j.gastro.2022.04.019

Mavuri, M., & Chakrabarty, S. (2024, December). Geospatial analysis of socioeconomic equity and environmental factors influencing lung cancer prevalence in the U.S. In 2024 IEEE International Conference on Bioinformatics and Biomedicine (BIBM) (pp. 6597–6604). IEEE. 10.1109/BIBM62325.2024.10822373

Megwalu, U. C. (2017). Impact of county-level socioeconomic status on oropharyngeal cancer survival in the United States. Otolaryngology–Head and Neck Surgery, 156(4), 665–670. 10.1177/0194599817691462

Montgomery, A., Vadapalli, R., Dinenno, F. A., Schilling, J., Jain, P., Jacob, A., Chism, D., & Shanker, A. (2025). Machine learning to evaluate the effects of non-clinical social determinant features in predicting colorectal Cancer mortality in a medically underserved Appalachian population. Scientific Reports, 15(1), 25781. 10.1038/s41598-025-11074-y

Niu, L., Hu, L., Li, Y., & Liu, B. (2022). Correlates of cancer prevalence across census tracts in the United States: A bayesian machine learning approach. Spatial and Spatio-temporal Epidemiology, 42, 100522. 10.1016/j.sste.2022.100522

Obeng-Gyasi, S., Obeng-Gyasi, B., & Tarver, W. (2021). Breast cancer disparities and the impact of geography. Surgical Oncology Clinics of North America, 31(1), 81. 10.1016/j.soc.2021.08.002

O’brien, R. M. (2007). A caution regarding rules of thumb for variance inflation factors. Quality & Quantity, 41(5), 673–690. 10.1007/s11135-006-9018-6

Palliyaguru, N., Chennamangalam, J., Liyanage, S., Wellalage, B. K. H., Arangala, C., Armstrong, N. M., & Palliyaguru, D. L. (2024). Geographical mapping of colorectal cancer incidence risk factors in the United States using statistical and machine learning approaches. Research Square. 10.21203/rs.3.rs-4752477/v1

Rahib, L., Wehner, M. R., Matrisian, L. M., & Nead, K. T. (2021). Estimated projection of US cancer incidence and death to 2040. JAMA Network Open, 4(4), e214708–e214708. 10.1001/jamanetworkopen.2021.4708

Ray, K., & Ghosh, R. (2025). Leveraging machine learning to analyze socioeconomic disparities in U.S. cancer clinical trials. In 2025 7th International Congress on Human-Computer Interaction, Optimization and Robotic Applications (ICHORA) (pp. 1–5). IEEE. 10.1109/ICHORA65333.2025.11017141

Robilotti, E. V., Babady, N. E., Mead, P. A., Rolling, T., Perez-Johnston, R., Bernardes, M., Bogler, Y., Caldararo, M., Figueroa, C. J., Glickman, M. S., Joanow, A., Kaltsas, A., Lee, Y. J., Morjaria, S., Nawar, T., Papanicolaou, G. A., Predmore, J., Redelman-Sidi, G., Schmidt, E., … Kamboj, M. (2020). Determinants of COVID-19 disease severity in patients with cancer. Nature Medicine, 26(8), 1218–1223. 10.1038/s41591-020-0979-0

Robinson, J. R. M., Beebe-Dimmer, J. L., Schwartz, A. G., Ruterbusch, J. J., Baird, T. E., Pandolfi, S. S., Hastert, T. A., Quinn, J. W., & Rundle, A. G. (2021). Neighborhood walkability and body mass index in African American cancer survivors: The Detroit research on cancer survivors study. Cancer, 127(24), 4687–4693. 10.1002/cncr.33869

Ruckthongsook, W., Tiwari, C., Oppong, J. R., & Natesan, P. (2018). Evaluation of threshold selection methods for adaptive kernel density estimation in disease mapping. International Journal of Health Geographics, 17(1), 10. 10.1186/s12942-018-0129-9

Salmeron, B., Mamudu, L., Liu, X., Whiteside, M., & Williams, F. (2021). Assessing health disparities in breast cancer incidence burden in Tennessee: Geospatial analysis. BMC Women’s Health, 21(1), 186. 10.3322/caac.21703

Shaik, F. (2024). Predictive modeling and spatial analysis of cervix uteri and breast cancer in India using machine learning and big data frameworks. Iranian Journal of Blood and Cancer, 16(4), 20–29.

Shao, K., & Feng, H. (2022). Racial and ethnic healthcare disparities in skin cancer in the United States: a review of existing inequities, contributing factors, and potential solutions. The Journal of Clinical and Aesthetic Dermatology, 15(7), 16–22. https://pubmed.ncbi.nlm.nih.gov/35942012/

Smith, S., Sakhamuri, S., Guidry, C. M., & Mustata Wilson, G. (2025). Social vulnerability and cancer risk from air toxins in Louisiana: A spatial analysis of environmental health disparities. Frontiers in Public Health, 13, 1601868. 10.3389/fpubh.2025.1601868

Soerensen, S. J. C., Lim, D. S., Montez Rath, M. E., Chertow, G. M., Chung, B. I., Rehkopf, D. H., & Leppert, J. T. (2025). Pesticides and prostate cancer incidence and mortality: An environment wide association study. Cancer, 131(1), e35572. 10.1002/cncr.35572

Sobti, R. C., Thakur, M., & Kaur, T. (2024). Cancer: Epidemiology, racial, and geographical disparities. In Molecular Biomarkers for Cancer Diagnosis and Therapy (pp. 31–52). Springer Nature Singapore. 10.1007/978-981-99-3746-2_3

Sotomayor, L. N., Cracknell, M. J., & Musk, R. (2023). Supervised machine learning for predicting and interpreting dynamic drivers of plantation forest productivity in northern Tasmania, Australia. Computers and Electronics in Agriculture, 209, 107804. 10.1016/j.compag.2023.107804

Sun, Y., Ao, Z., Jia, W., Chen, Y., & Xu, K. (2021). A geographically weighted deep neural network model for research on the spatial distribution of the down dead wood volume in Liangshui National Nature Reserve (China). IForest-Biogeosciences and Forestry, 14(4), 353. 10.3832/ifor3705-014

Tabassum, N., Chowdhury, M. M., McMahan, C. S., Self, S., Isanovic, M., Correa-Velez, K., Sellers, S. C., Norman, R. S., & Rennert, L. (2025). Granular insights: A wastewater-based machine learning approach for localized COVID-19 hospitalization forecasting. medRxiv. 10.1101/2025.06.25.25330294

Tailor, T. D., Choudhury, K. R., Tong, B. C., Christensen, J. D., Sosa, J. A., & Rubin, G. D. (2019). Geographic access to CT for lung cancer screening: A census tract-level analysis of cigarette smoking in the United States and driving distance to a CT facility. Journal of the American College of Radiology, 16(1), 15–23. 10.1016/j.jacr.2018.07.007

Tesfaw, L. M., & Muluneh, E. K. (2020). Modeling the spatial distribution of cancer and determining the associated risk factors. Cancer Informatics, 19. 10.1177/1176935120939898

Vatcheva, K. P., Lee, M., McCormick, J. B., & Rahbar, M. H. (2016). Multicollinearity in regression analyses conducted in epidemiologic studies. *Epidemiology (Sunnyvale*, Calif.), 6(2), 227. 10.4172/2161-1165.1000227

Vieira, V. M., VoPham, T., Bertrand, K. A., James, P., DuPré, N., Tamimi, R. M., Laden, F., & Hart, J. E. (2020). Contribution of socioeconomic and environmental factors to geographic disparities in breast cancer risk in the Nurses’ Health Study II. Environmental Epidemiology, 4(1), e080. 10.1097/EE9.0000000000000080

Wiese, D., DuBois, T. D., Sorice, K. A., Fang, C. Y., Ragin, C., Daly, M., Reese, A. C., Henry, K. A., & Lynch, S. M. (2024). An exploratory analysis of the impact of area-level exposome on geographic disparities in aggressive prostate cancer. Scientific Reports, 14(1), 16900. 10.1038/s41598-024-63726-0

Zhang, S., Jin, J., Zheng, Q., & Wang, Z. (2025). Building a cancer risk and survival prediction model based on social determinants of health combined with machine learning: A NHANES 1999 to 2018 retrospective cohort study. Medicine, 104(6), e41370. 10.1097/MD.0000000000041370

